# Cross-species Multimodal Single-cell Profiling of Dorsal and Ventral Prefrontal Cortex Reveals Cell-type Divergence and PTSD-associated Regulatory Landscapes

**DOI:** 10.1101/2025.07.28.25332052

**Authors:** Feiyang Zhang, Kaixin Huang, Zechen Liu, Xin Liu, Qiongyi Zhao, Xinyan Li, Wei Wei, Xiang Li

## Abstract

The prefrontal cortex subregions—particularly the prelimbic (PL) and infralimbic (IL) cortices in rodents and dorsal anterior cingulate (dACC) and ventromedial prefrontal cortex (vmPFC) in humans—exhibit functionally specialized yet interconnected roles in PTSD pathogenesis. While PL/dACC are implicated in fear acquisition, IL/vmPFC are associated with fear extinction. However, the inherent difference and cross-species molecular signatures underlying these functional parallels remain unresolved. To bridge the gap, we integrate single-nucleus RNA-seq, ATAC-seq, and spatial transcriptomics across mouse PL/IL and human dACC/vmPFC to construct a cross-species, multi-omic atlas. We delineate conserved/divergent gene regulatory networks (GRNs), with emphasis on excitatory neuron evolution. By incorporating PTSD GWAS data and gene expression changes from vmPFC of PTSD patients, we identify cell-type-specific PTSD risk enrichment, SNP-anchored GRNs linked to PTSD heritability, and stress-primed chromatin states in candidate neurons. This work provides a molecularly resolved subregional atlas and advances translational understanding of PTSD-related gene regulation divergence and complement the present multi-omic research of PTSD.

## Introduction

The prelimbic cortex (PLPFC) and infralimbic cortex (ILPFC) cortices in rodents, along with their putative human homologs—the dorsal anterior cingulate cortex (dACC) and ventromedial prefrontal cortex (vmPFC)—exhibit striking functional divergence in fear regulation and PTSD pathogenesis. In rodents, the PLPFC promotes fear expression and threat response, while the ILPFC is critical for fear extinction and behavioral flexibility (refs^1,2^). Similarly, in humans, dACC hyperactivity correlates with exaggerated fear recall and symptom severity in PTSD patients, whereas vmPFC hypoactivity predicts impaired extinction retention (ref^3,4,5,6^). These observations suggest conserved functional specialization across species, where PL/dACC drive fear acquisition and IL/vmPFC mediate extinction.

Despite extensive functional characterization of PL/ILPFC and dACC/vmPFC in fear processing or in other brain regions (refs^7,8,9^), the cell-type-specific molecular profiles and regulatory divergence remain systematically uncharacterized, notably, no study has comprehensively compared: (1) the transcriptional and epigenetic signatures of cell subpopulations between PL/dACC versus ILPFC/vmPFC, or how stress-induced chromatin remodeling differs across these regions.

Meanwhile, preclinical PTSD studies rely heavily on rodent fear conditioning paradigms face challenges in fully capturing the complex, heterogeneous nature of PTSD (refs^10,11^). Preclinical PTSD studies using rodent fear conditioning paradigms have limited translation to human trials, requiring conceptual and technological improvements (ref^12^). A critical barrier is the lack of molecularly defined cross-species maps aligning functional homology with cell-type-specific gene regulation. Rodent PFC and human PFC share similar connectivity patterns (ref^13^), but transcriptional and cytoarchitectural differences suggest evolutionary divergence (refs^14,15^). Bridging this rodent-to-human knowledge gap is clinically imperative, which could transform translational psychiatry by pinpointing conserved PTSD-relevant circuits and druggable targets.

Recent advances in single-nucleus RNA sequencing (snRNA-seq) and single-nucleus ATAC-seq (snATAC-seq) have revolutionized our ability to dissect cell-type-specific gene expression and chromatin accessibility at unprecedented resolution. snRNA-seq profiles transcriptomes from individual nuclei, enabling the identification of rare neuronal subtypes and their transcriptional states (ref^16^). Complementary to this, snATAC-seq maps open chromatin regions, revealing cis-regulatory elements (CREs) such as enhancers and promoters, which are critical for understanding gene regulation dynamics. The integration of these datasets allows reconstruction of gene regulatory networks (GRNs), linking transcription factors (TFs) to their target genes through motif analysis and co-accessibility profiling. Spatial transcriptomics further bridges molecular profiles with anatomical context, resolving regional heterogeneity in complex tissues like the brain. Techniques such as Visium (10x Genomics) have been pivotal in mapping gene expression gradients across cortical layers and subregions (e.g., PLPFC vs. ILPFC), offering insights into how spatial organization influences functional specialization.

Chromatin-primed genes (CPGs) are defined as loci that reside in a transcriptionally permissive but inactive state, marked by accessible chromatin (e.g., ATAC-seq peaks) and histone modifications (e.g., H3K4me1 at enhancers), yet require external stimuli (e.g., stress) for full activation (ref^17^). These genes are hypothesized to mediate rapid transcriptional responses in cells upon environmental cues such as diseases (refs^18,19,20^). snATAC-seq can identify primed loci by detecting stress-inducible CPGs that gain accessibility post-stimulus, while snRNA-seq reveals their downstream transcriptional cascades.

While single-nucleus multi-omics approaches (snRNA-seq, snATAC-seq, and spatial transcriptomics) have advanced our understanding of PTSD-related prefrontal dysfunction, prior studies have largely focused on *either* rodent vmPFC *or* human PFC independently, leaving their cross-regional and cross-species molecular alignment unresolved. Existing reaserches have yet to resolve whether PL/dACC and IL/vmPFC are distinguished by fundamentally distinct gene regulatory networks (GRNs) or share a core transcriptional program and integrate the difference with the PTSD disease model. Also, the evolutionary conservation of stress-response pathways in homologous cell types in the respective region remains equally unclear. Furthermore, while epigenetic mechanisms play a crucial role in fear conditioning and extinction, with implications for treating PTSD, such as chromatin-modifying enzymes p300/CBP-associated factor (PCAF), regulate gene expression associated with fear extinction memory formation (ref^21,22,23^), chromatin-primed genes (CPGs) - pre-marked by high accessibility but requiring stress for activation - have not been comprehensively identified in the sub-regions in PFC, the regional specificity and contribution to PTSD still remain unexplored.

Our study addresses these gaps through 1) cross-species, cross-regional multi-omics integration with PTSD disease model, 2) defining the transcriptomic, epitranscriptomic (chromatin accessibility) and GRN architecture that differentiates the two brain regions at single-cell resolution, 3) finding conserved and divergent gene expression pattern using PTSD GWAS SNPs and stress-responsive CPGs to predictict PTSD susceptibility cell types and genes. Thus provide a comprehensive cross species reference atlas and susceptibility map for PTSD research.

## Results

### Gene and Chromatin Accessibility Divergence Revealed by Single-Nucleus Multi-Omic Profiling for Mouse

To resolve molecular distinctions between mouse prelimbic (PL) and infralimbic (IL) prefrontal cortex, we micro-dissected these subregions under naïve conditions (Fig. 1a) and performed parallel snRNA-seq and snATAC-seq using established droplet-based protocols. After standard preprocessing and quality control (Methods), snRNA-seq of 14,494 nuclei identified 7 excitatory neuron subtypes, 6 inhibitory neuron populations, and non-neuronal cells (Fig. 1b), with canonical markers confirming cluster identities (*Slc17a7*+ excitatory neurons, *Gad1*+ inhibitory neurons; Extended Data Fig. 1a-c). Excitatory neurons constituted 64.98% of all nuclei (Fig. 1c). Visualization by log□ differential density revealed preferential enrichment of excitatory neurons in PL compared to IL (Fig. 1d). snATAC-seq of 11,282 nuclei recapitulated major clusters observed in transcriptomic data (Fig. 1e), with excitatory neurons again dominating (77.62% of nuclei; Fig. 1f). Chromatin accessibility-based UMAP projections mirrored the PL-enriched distribution pattern seen in snRNA-seq (Fig. 1g). Canonical markers could be found from Azimuth resources (ref^24^). Meanwhile, we also applied MultiK (ref^25^) to find more sub-clusters from major cell types. We further found marker genes and features for sub-types (ref^26^) (Extended Data Fig. 1d-e). We then co-embedded snRNA-seq and snATAC-seq using scBridge (ref^27^) (Extended Data Fig. 1f). Systematic quantification of regional cell-type distribution bias confirmed consistent neuronal localization across both assays, though glial populations exhibited batch-dependent variation (Fig. 1h).

**Figure 1.**
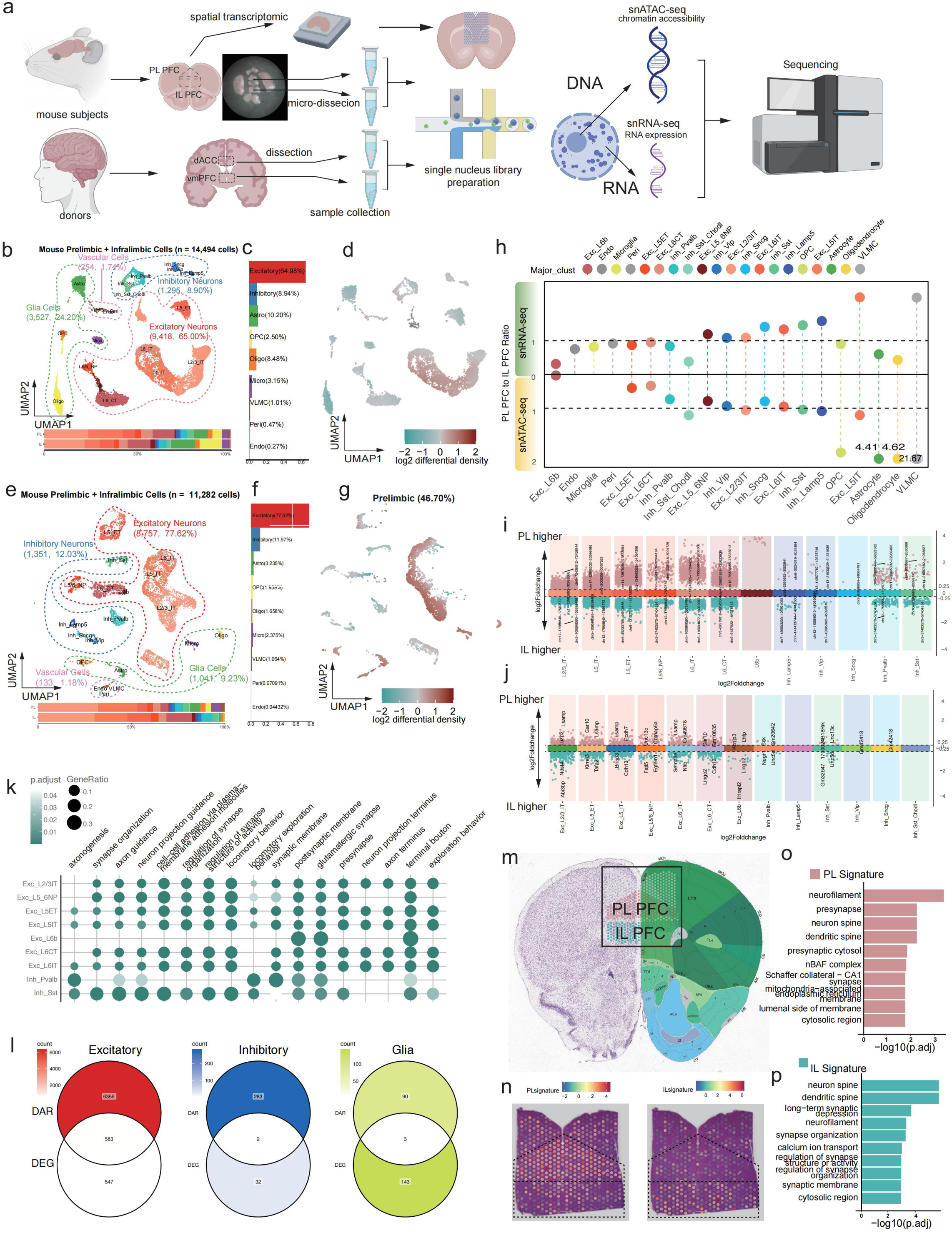
Single-cell multi-omic profiling of mouse prefrontal subregions. Integrates gene expression, chromatin accessibility, regulatory relationships, and spatial transcriptomics. a, Schematic of the experimental workflow. The study utilized both mouse and human subjects. Mouse prelimbic (PL) and infralimbic (IL) prefrontal cortex (PFC) subregions were microdissected, collected separately, and subjected to snRNA-seq and snATAC-seq. A coronal section encompassing both PL and IL was paraffin-embedded, sectioned, and used for spatial transcriptomics. For human donors, dorsal anterior cingulate cortex (dACC) and ventromedial PFC (vmPFC) tissues were collected from each individual. All human and mouse samples underwent 10x Genomics single-cell library preparation followed by snRNA-seq and snATAC-seq. b, UMAP projection of mouse snRNA-seq data. Cell types were annotated, revealing 7 excitatory neuron types, 5 inhibitory interneuron types, and 4 glial cell types. Bar plots below show the proportion of each major cell type within the PL and IL subregions. c, Proportions of major cell types identified in mouse snRNA-seq. d, UMAP colored by the relative enrichment of cells in PL versus IL subregions. e-g, UMAP projection (e), cell type proportions (f), and UMAP colored by relative enrichment in IL versus PL (g) for mouse snATAC-seq data. h, Relative distribution of each major cell type between PL and IL subregions in snRNA-seq (top) and snATAC-seq (bottom). X-axis: Major cell types; Y-axis: Ratio of cell type proportion in PL to that in IL. Dashed line indicates equal proportion (ratio=1). i, Identification of differentially expressed genes (DEGs) between PL and IL within each cell type. Points represent genes; X-axis incorporates random jitter within each cell type; Y-axis: log2 fold change (FC) of expression in PL relative to IL. j, Bubble plot of Gene Ontology (GO) enrichment for DEGs identified in panel i. X-axis: GO terms; Y-axis: Major cell types; Color: Adjusted p-value; Size: Gene Ratio (percentage of DEGs in the cell type associated with the GO term). k-l, Top regulon sets by regulon specificity score (RSS) identified through SCENIC+ integrative analysis of snATAC-seq and snRNA-seq. k, Activator regulons. l, Repressor regulons. For each cell in the heatmap: Upper left triangle - Expression level of regulon target genes; Lower right triangle - Chromatin accessibility level of transcription factor (TF)-bound regions. m, Example of an activator regulon: Klf10 binds an open region upstream of *Fndc9* (functioning as an enhancer), promoting higher expression of *Fndc9* in PL within the excitatory L2/3IT neuron type. n, Schematic of the mouse spatial transcriptomics region (black box). Boundaries between PL and IL were defined using histological staining from the Allen Brain Atlas. o-q, Identification and functional annotation of spatial gene expression signatures specific to PL (o), IL (p), and their comparison (q).

Differential gene expression and chromatin accessibility analyses discovered pronounced subregional divergence. Excitatory neurons displayed significantly more differentially accessible peaks (DARs) and differentially expressed genes (DEG) between PL and IL than inhibitory neurons (Fig. 1i-j). For DEGs, functions are enriched for axon guidance and neuronal projection pathways (Fig. 1k). Surprisingly, DARs exhibited substantial overlap with DEGs, particularly in regulatory elements linked to neuronal morphogenesis in excitatory neuron, but less overlap in inhibitory neurons and glia cells, suggesting a higher diverse in epigenetic regulation in inhibitory neurons and glia cells in different brain regions (Fig. 1l). Spatial transcriptomics independently validated the difference of ILPFC and PLPFC. Region-specific signatures—PL-enriched for neurofilament assembly genes, IL-enriched for synaptic genes (Fig. 1m-p)—aligned with functional annotations from snRNA-seq. Collectively, these results demonstrate that while PL and IL share broad cellular composition, they exhibit pervasive differences in transcriptional and epigenetic landscapes, particularly within excitatory neurons, reflecting inherent functional specialization.

### Gene and Chromatin Accessibility Divergence Revealed by Single-Nucleus Multi-Omic Profiling for Human

To delineate human-specific molecular profiles, we analyzed snRNA-seq and snATAC-seq data from dissected dACC and vmPFC subregions across 20 postmortem tissues (Methods), generating comprehensive transcriptomic (n = 203,487 nuclei) and chromatin accessibility (n = 211,592 nuclei) profiles (Fig. 2a). We leveraged RPCA from Seurat and Signac (ref^28–30)^ to transfer annotation labels from mouse to human, with canonical marker genes for validations (ref^31^) (Extended Data Fig. 2a-c). Same as in mouse, we similarly found sub-clusters using MultiK (ref^32^) and profiled the marker genes for each sub-cluster (Extended Data Fig. 2d-e). Consistent with established human cortical organization (ref^33^), oligodendrocytes constituted the predominant cell population (54.48% of nuclei; Fig. 2b,c), contrasting sharply with murine observations. snATAC-seq confirmed comparable cellular composition (Fig. 2d-f), with minimal global differences between vmPFC and dACC. Specifically, comparing to mouse, we didn’t find endothelia cells being identified in the human PFC.

**Figure 2.**
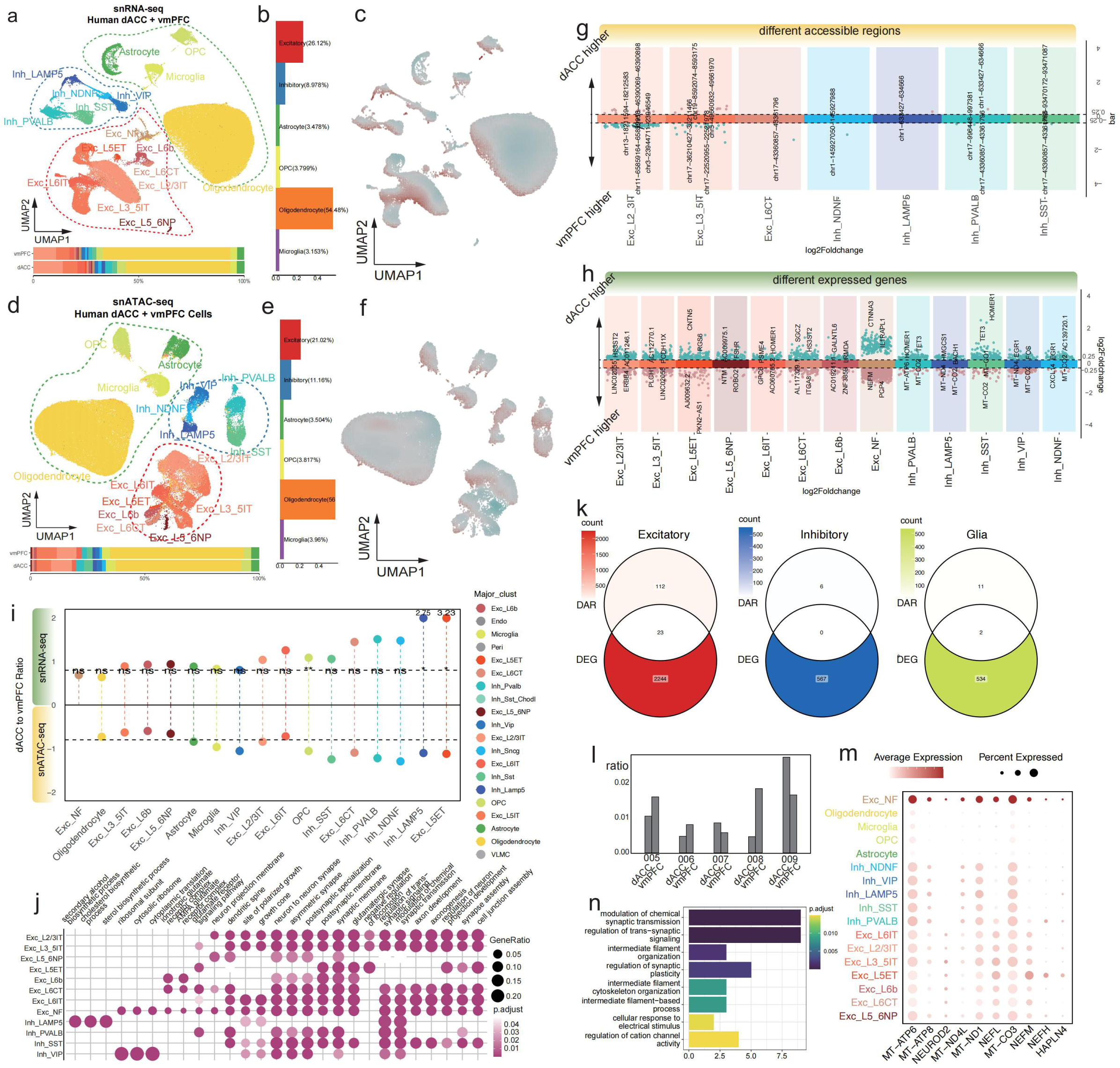
Single-cell multi-omic profiling of human cortical subregions. Integrates gene expression, chromatin accessibility, and regulatory relationships. a, Integrated UMAP projection of snRNA-seq data from all human samples across dACC and vmPFC subregions. Cell types annotated include 8 excitatory neuron types, 5 inhibitory interneuron types, and 4 glial cell types. Bar plots below show the proportion of each major cell type within vmPFC and dACC. b, Cell type proportions across all donors and subregions. c, UMAP colored by the relative enrichment of cells in dACC versus vmPFC. d-f, Integrated UMAP projection (d), cell type proportions (e), and UMAP colored by relative enrichment in dACC versus vmPFC (f) for human snATAC-seq data. g, Relative distribution of each major cell type between dACC and vmPFC in snRNA-seq (top) and snATAC-seq (bottom). Format as in Fig. 1h. Dashed line indicates equal proportion. h, Identification of DEGs between dACC and vmPFC within each cell type. Format as in Fig. 1i. i, Bubble plot of GO enrichment for DEGs identified in panel h. Format as in Fig. 1j. j-k, Top regulon sets by RSS from SCENIC+ analysis. j, Activator regulons. k, Repressor regulons. Heatmap format as in Fig. 1k-l. l, Example activator regulon: FOS, a neuronal immediate early gene, binds an open region downstream of *RPTOR* (functioning as an enhancer), activating higher expression in vmPFC within the excitatory L6 CT neuron type. m, Proportion of the excitatory neuron subtype Exc-NF within each donor’s dACC and vmPFC. Donor IDs: 005-009. n, Bubble plot showing expression of Exc-NF marker genes within the Exc-NF subtype and across all other cell types. Color: Mean expression level; Size: Fraction of cells expressing the gene. o, Functional annotation of marker genes for the Exc-NF neuronal subtype.

Differential expression (|log□FC| > 0.25, FDR < 0.05) and chromatin accessibility (|log□FC| > 0.25, FDR < 0.05) analyses demonstrated pronounced subregional divergence in both excitatory and inhibitory neuronal populations (Fig. 2g-h). Comparing to mouse, the overlap of DEG and DAR decreased in excitatory neurons, and each cell-type has a potentially more diverse epigenetic resolution within brain regions in human.

Meanwhile, we found that microglia, inhibitory neuron LAMP5 and Excitatory neuron L5ET with a significant higher enrichment in dACC cortex (Fig. 2i). To find function of different expression genes, we performed GO enrichment (ref^34^). Unlike murine patterns, human inhibitory neurons exhibited substantial transcriptional heterogeneity, with LAMP5+ neurons showing metabolic pathway enrichment and VIP+ neurons displaying ribosome related signatures. Excitatory neurons maintained conserved functional annotations in synaptic transmission and dendritic morphogenesis, similar to murine findings.

Notably, we identified a novel excitatory neuronal state (Exc_NF) conserved across all donors (Fig. 2m), defined by co-expression of neurofilament genes (*NEFL*, *NEFM*, *NEFH*), which are essential cytoskeletal components in neurons (refs^35^). We also found the neurogenic transcription factor *NEUROD2* and mitochondrial transcripts as its markers (Fig. 2n). Functional annotation revealed enrichment in electrochemical transmission and voltage-gated channel activity (Fig. 2j). Integration with snATAC-seq data shows a close relationship with Oligodendrocyte (Extended Data Fig. 2f), which fits the function of neurofilament in providing structural support for neurons, particularly in myelinated axons (ref^36^). This transcriptional signature suggests potential neurofilament-mediated mitochondrial recruitment—a mechanism previously implicated in axonal energy homeostasis (refs^37,38^), but not previously observed in cortical somata.

A brief cross-species comparisons revealed greater divergence in inhibitory neuron regulatory landscapes than excitatory populations, with limited conservation of canonical marker genes. We found that very interestingly, *Lamp* and *Ndnf* have co-expression in mouse prefrontal cortex is having, but never co-expressed in human prefrontal cortex for human gene *LAMP* and *NDNF*, revealing the diverse expression pattern in human and mouse inhibitory neuron (Fig. S2).

These results reveal pronounced subregional regulatory divergence—particularly in inhibitory neurons comparing to murine—and demonstrate fundamental human-specific reorganizations in prefrontal cytoarchitecture and gene regulatory programs, contrasting with murine patterns while revealing evolutionarily conserved stress-response principles in excitatory neurons.

### Gene Regulation Network Reveals the Regulation Pattern Cross Regions and Species

To identify cross-region and cross-species gene regulatory networks, we applied SCENIC+ (ref^39^)—an integrative single-cell multi-omic framework that infers enhancer-linked regulons (eRegulons, eRg) using combined chromatin accessibility (scATAC-seq) and expression data (scRNA-seq). We detected a total of 26,882 enhancer-based regulons in mouse PLPFC and ILPFC, 17,884 in human dACC and vmPFC. To assign each eRg to a cell-type or region, we leveraged RSS score found by SCENIC+ (Extended Data Fig. c-d). Furthermore, we annotated their upstream/downstream enhancer binding regions and target genes against whether it’s defined as a marker for certain cell-type (Methods), retaining only eRgs with congruent region and marker annotations as cell-type specific eRgs regulated by multiple TFs (Extended Data Fig. 3a-b). This yielded 37 cell- and region-specific transcription factors comprising 909 eRgs, including notable TFs such as Klf10, which is related with neuron development and highly conserved in human and mouse (ref^40,41,42^) and Neurod2, which coordinates synaptic innervation and intrinsic excitability to balance excitation and inhibition in cortical pyramidal neurons (ref^43^) (Fig. 3a–b). Within mouse PL, the Sncg+ inhibitory neuron cluster displayed the highest number of cell-type-specific eRgs. Several region-specific eRgs overlapped with differentially expressed genes identified earlier, for instance, we found that Klf10’s enrichment in Exc_L2/3IT neuron, regulates Fndc9, a gene classified to having interactions with some miRNAs (refs^44^), which is a significantly higher expressed in PLPFC (Extended Data Fig. 3e). Meanwhile, we found FOS (refs^45^), a well-known immediate early gene, which regulate RPTOR, a highly methylated gene reported closely related with PTSD (refs^46,47^), is higher expressed in human vmPFC (Extended Data Fig. 3f).

**Figure 3.**
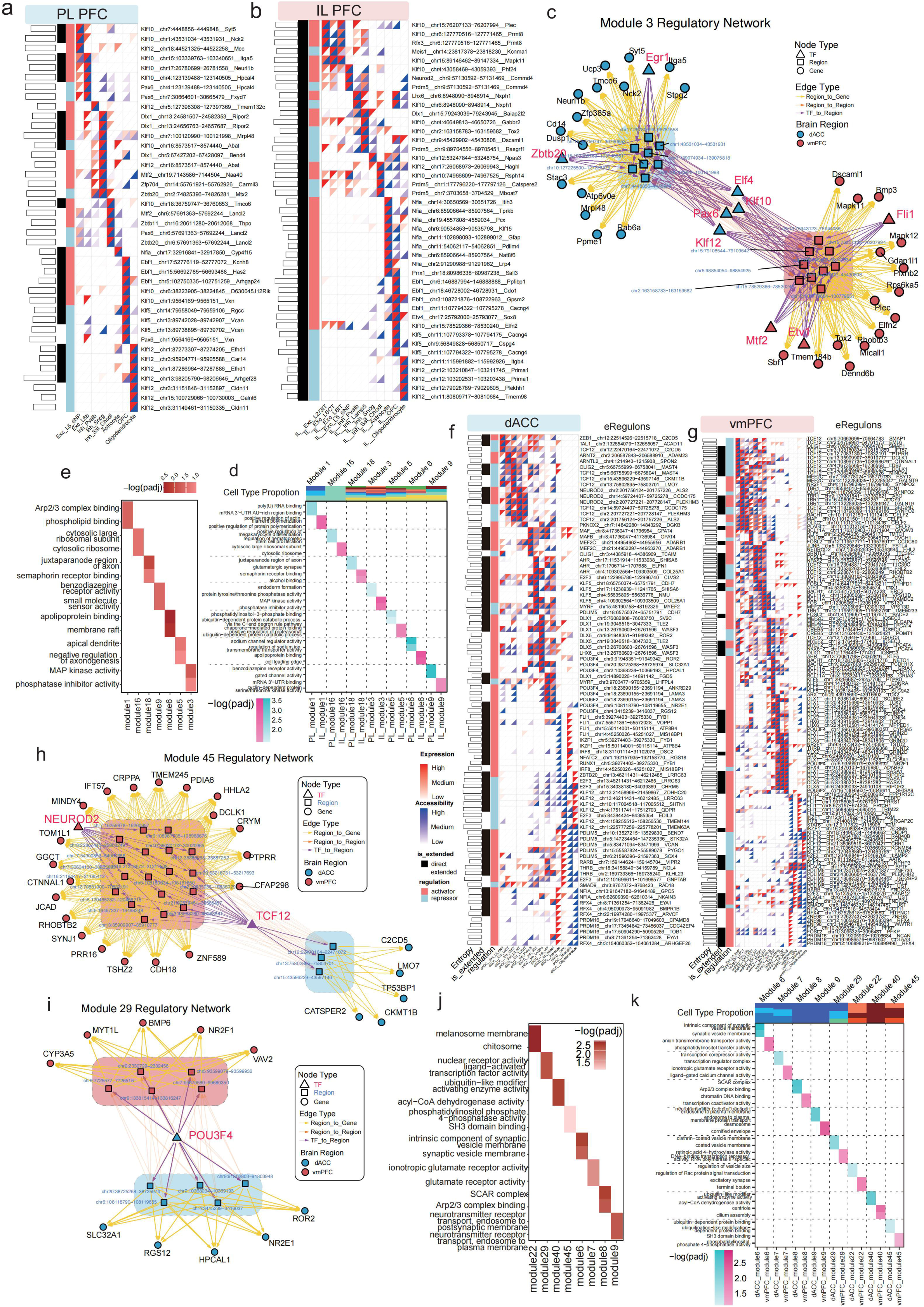
Regulatory network analysis in mouse and human. a-b, Heatmaps of high-scoring cell-type-specific enhancer regulons (eRGs) identified in mouse PL (a) and IL (b). Left bar plot: Information entropy per eRG. Black marks: SCENIC+-identified direct eRGs; Red/Blue marks: Activator/Repressor. Upper left triangle: Expression level of regulated genes; Lower right triangle: Chromatin accessibility of TF-bound regions. c, Detailed regulatory network for mouse module 3 in PL and IL. Triangles: TFs; Squares: TF-bound regions; Circles: Target genes. Blue and red boxes highlight region sets correlating within PL and IL networks, respectively. d-e, Functional annotation (GO analysis) of genes regulated by neuron-associated modules, analyzed separately for PL (d) and IL (e). f-g, Heatmaps of high-scoring cell-type-specific eRGs identified in human dACC (f) and vmPFC (g). Format as in a-b. h-i, Detailed regulatory networks for human module 45 (h) and module 29 (i) in dACC and vmPFC. Format as in c. j-k, Functional annotation (GO analysis) of genes regulated by neuron-associated modules, analyzed separately for dACC (j) and vmPFC (k).

To assess gene regulatory network (GRN) divergence, for each pair of subregions (intra- or inter-species), we computed a similarity score based on shared TF-gene connections (Methods). Links with similarity >□0.5 were assembled into regulatory modules—clusters of TFs, regions, and target genes with no cross-module connections. We identified 19 modules in mouse PL and ILPFC. Modules comprised genes from multiple cell types: for instance, Module 3 was enriched in excitatory neuron genes, whereas Module 1 was dominated by inhibitory neuron targets (Extended Data Fig. 4a). Notably, several modules were composed exclusively of glial cell eRgs, indicating substantial regulatory divergence for glia between IL and PL. The regulation in modules reveals that there are high overlap of TFs through a set of connected regions from one brain region to another. Functional profiling—both pooled and region-specific (Fig. 3e)—revealed striking regional specialization: in Module 3, PL genes were associated with protein phosphatase activity and endodermal differentiation, whereas IL genes were enriched for MAP kinase signaling.

Applying the same analytical pipeline to human dACC and vmPFC, we identified a greater number of cell- and region-specific eRgs—3,735 eRgs regulated by 111 unique transcription factors (Extended Data Fig. 3b)—indicating a broader regulatory network. From all GRN, following the same pipeline, we also uncovered 45 regulatory modules in the human PFC, including several composed entirely of oligodendrocyte eRgs, reflecting the increased complexity of human glia function (Extended Data Fig. 4b). There are also other modules were strictly excitatory (e.g., modules 22, 40, 45) or inhibitory (modules 6–9), signifying cell-type-specific regulatory partitioning.

### Similarity and Divergency from Mouse to Human from the Perspective of Homologs

To investigate cross-species homology, we conducted integrated analysis of homologous cell types and orthologous genes between mouse and human. Co-embedding of single-nucleus profiles demonstrated strong alignment of homologous cell populations across species (Fig.□4a, S4). Label transfer (Methods) analysis measuring all features further confirmed high correspondence among matched cell types, revealed the overall similarity of homolog cell types from mouse to human. Notably, the mouse *Sncg*LJ inhibitory neuron cluster exhibited strong similarity with human *VIP*LJ and *NDNF*LJ inhibitory neurons, consistent with prior cross-species comparisons of interneuron lineages. Given that transcription factors (TFs) serve as developmental markers and drivers of cell identity, we quantified TF expression similarity between homologous cell types. Across evaluated TFs, mouse and human homologs showed robust concordance in expression profiles, underscoring conserved regulatory control.

We then quantified similarity using Jaccard Index (JI) based on robust marker genes (Fig.□4d) and cell-type–specific chromatin peaks (Fig.□4e). When pooling across cortex regions (IL□+□PL and dACC□+□vmPFC), glial cell types displayed the highest cross-species similarity, followed by inhibitory neurons, with excitatory neurons occupying an intermediate position (Fig.□4d–e). Clusters 56NP, 5ET, and 6b demonstrated strong interspecies similarity based on marker gene overlap (upper panel). However, incorporating region-specific markers reduced JI across PL-dACC subtypes—particularly for neuronal populations except 2/3IT (middle panel). In contrast, between IL and vmPFC, excitatory subtypes dL6b and CT maintained elevated similarity (lower panel).

Orthologous genes were further assessed using Divergence Score (DS) (ref^33^), with |DS|□>□2 denoting high divergence (Methods). We categorized genes into housekeeping, robust markers, DEGs, and eRg targets, and computed DS distributions per major cell-type (Fig.□4f, Extended Data Fig. 5a), color-coded by gene category (Methods). Most orthologs exhibited low divergence, though a subset showed pronounced interspecies differences. Genes with high DS in human were predominantly human-derived markers or DEGs (and vice versa). Functional enrichment of highly divergent genes in excitatory and inhibitory neurons revealed synapse-related, migration, and cell junction pathways. Surprisingly, divergent glial genes also implicated neuron-centric processes, such as synapse organization, axonogenesis, and migration (Fig. 4g-i).

**Figure 4.**
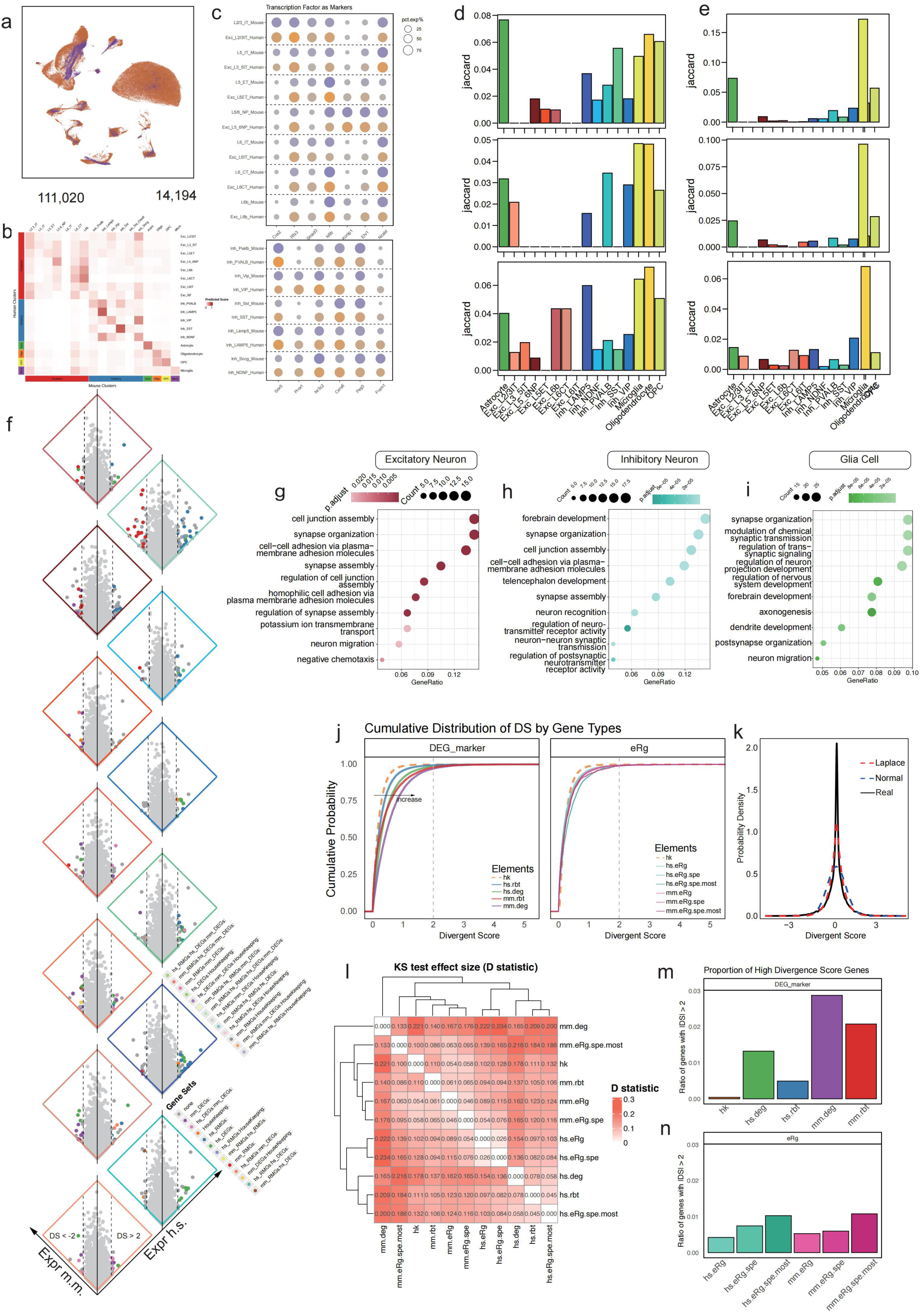
Analysis of cell type homology and gene expression divergence between mouse and human. a, Heatmap depicting similarity between homologous mouse (x-axis) and human (y-axis) major cell types. b-c, Jaccard index (JI) assessing similarity using marker genes (b) or marker chromatin accessibility peaks (peaks) (c) identified by starTracer. Upper panel: JI comparing pooled dACC/vmPFC to pooled PL/IL; Middle panel: PL vs dACC; Lower panel: IL vs vmPFC. Higher JI indicates greater similarity. d, Divergence in expression levels of conserved genes within homologous cell types. Y-axis (left): Log1p expression in mouse; X-axis (right): Log1p expression in human. Dashed lines indicate Deviation Score (DS) = ±2. Genes outside these lines exhibit significant expression divergence. Genes are categorized into five elements: Housekeeping genes, Mouse marker genes, Mouse PL/IL DEGs, Human marker genes, Human dACC/vmPFC DEGs. Color denotes the combination of elements assigned to each gene. e-g, Bubble plots of GO enrichment for genes with |DS| > 2 within each cell type. DEGs from excitatory and inhibitory neurons were annotated together (e), as were those from the four glial cell types (f). Format as in Fig. 1j. g, (Implied consolidation of e-f results for presentation). h, Left: Cumulative distribution functions (CDFs) of DS for each element type. Right: CDFs of DS for genes regulated by regulons (eRGs) classified by specificity: Non-specific (eRg), Cell-type-specific (eRg.Spe), and highly specific/Most specific (eRg.Spe.Most). i, Distribution of DS values closely follow a Laplacian distribution rather than a normal distribution. j, Kolmogorov-Smirnov (K-S) test results comparing DS distributions across element types and eRG specificity classes. Due to universally significant p-values (p<<0.05), the effect size (D statistic) is shown. Larger D indicates greater distribution difference. k-l, Bar plots showing the proportion of highly divergent genes (|DS| > 2) among the different elements (k) and among genes regulated by eRGs of varying specificity levels (l).

To integrate divergence with previously found regional different features and markers, as well as eRgs at different specificity level, we generated cumulative distribution curves ranking genes by DS and fitted them statistically for genes with correspondent labels (Fig.□4j–k, Methods). Housekeeping genes exhibited the lowest DS, followed sequentially by human robust markers, human DEGs, mouse robust markers, and mouse DEGs. Similarly, target genes of cell-type–specific eRegulons exhibited higher DS than housekeeping genes, indicating increased divergence. Remarkably, DS distributions closely followed Laplace curves across major cell types (Fig.□4k, Extended Data Fig. 5b), with Kolmogorov–Smirnov tests confirming significant distinctions between each gene category and eRegulon sets (Fig.□4l). Moreover, the proportion of highly divergent genes was lowest in housekeeping categories and increased progressively through markers, DEGs, and especially in cell-type-specific eRg targets (Fig.□4m–n). These findings corroborate our earlier observations: housekeeping genes are highly conserved across species, whereas mouse DEGs and markers are more divergent than their human counterparts, and genes regulated within highly cell-type-specific eRegulons exhibit the greatest cross-species divergence—suggesting a pivotal role for cell-type-specific regulatory complexity in driving species-specific traits.

#### Trajectory Analysis of L2/3IT Neuron Reveals Modules Related with Evolution from PL PFC to dACC

Excitatory L2/3 IT neurons, with its projection to multiple regions related with PTSD [cite], we investigated the trajectory of it from mouse to human to uncover the molecular mechanism driving cell heterogeneity cross-species (Fig. 5a). Sub-clusters of Excitatory L2/3 IT neurons were found by MultiK (ref^32^), giving 24 sub-clusters marked by a set of high specific genes (Methods). We used Monocle3 (ref^48^) to uncover the trajectories based on the cell similarity. Among multiple starting sites, we selected a trajectory representing the transforming from mouse nuclei to human nuclei (R^2 =0.245, p < 0.001) along the pseudo-time bin (Fig. 5c). Genes which vary along the trajectory were found and grouped as modules, then module score for each sub-cluster were profiled (Fig. 5b, Extended Data Fig. 6c). Based on the biological meaning of trajectory, modules explain the gradient from human to mouse as the proportion of human nuclei decrease.

**Figure 5.**
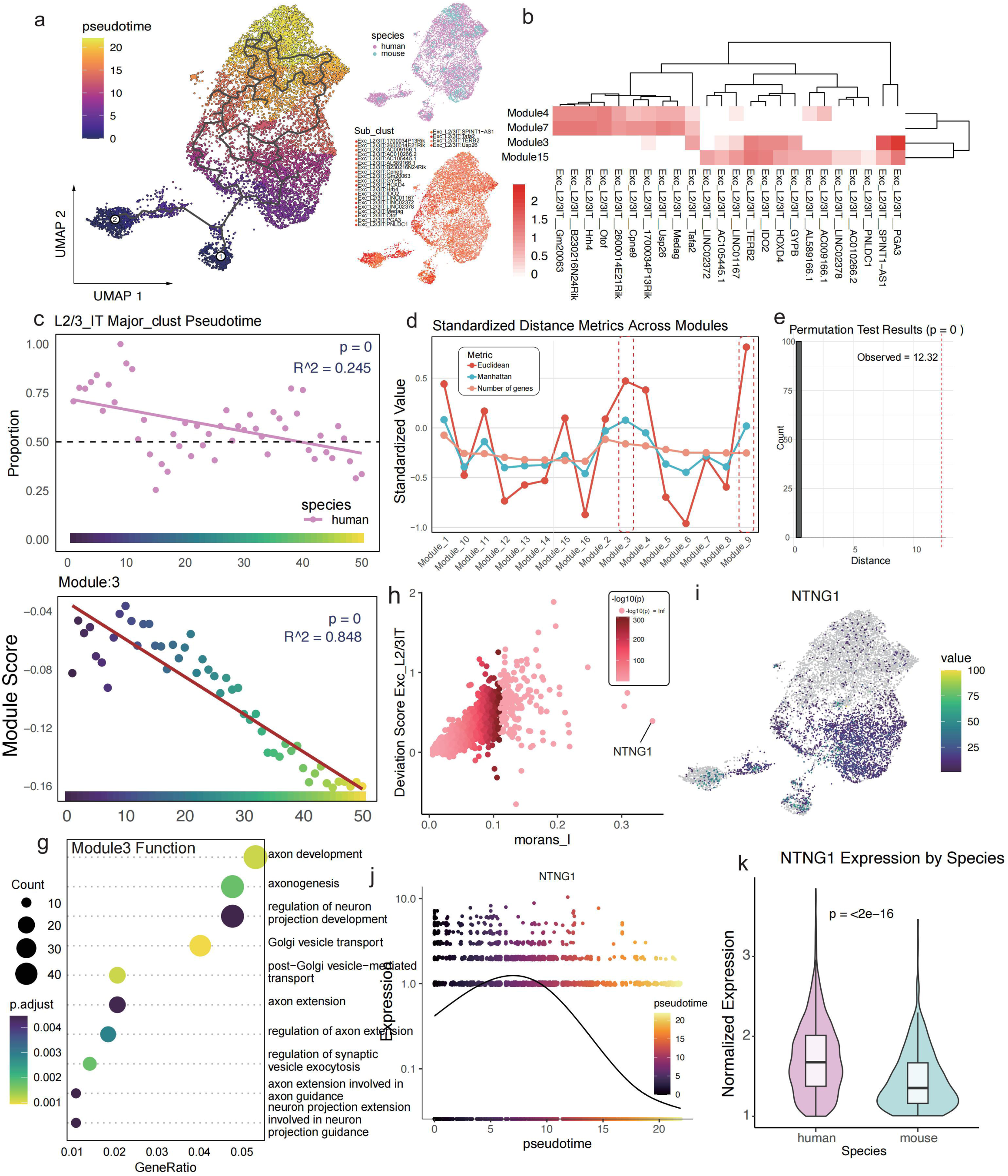
Trajectory analysis of L2/3IT excitatory neurons from mouse PL to human dACC. a, UMAP of integrated mouse and human L2/3IT neurons, colored by pseudotime inferred from defined Start Points 1 & 2. Top right: Species origin (human/mouse). Bottom right: L2/3IT subclusters identified by MultiK. b, Module activity scores derived from Non-negative Matrix Factorization (NMF) of trajectory DEGs across L2/3IT subclusters. c, Proportion of human cells within each of 50 pseudotime bins. Red line: Linear fit. d. Change in scaled Euclidean and Manhattan distances between human and mouse cells in the high-dimensional space upon sequential removal of each NMF module. Red dashed box highlights the module whose removal maximally increases distance. e, Permutation test p-value (p=0) for the effect of module 3 removal (from d). f, Module 3 activity score across pseudotime bins. Red line: Linear fit. g, Bubble plot of GO enrichment for genes within module 3. h, Scatter plot of spatial distribution metric (Moran’s I) for trajectory DEGs versus their DS. X-axis: Moran’s I; Y-axis: DS. Positive correlation is observed. i, Feature UMAP plot showing expression of *STXBP6* (a module 3 gene) in L2/3IT neurons. j-k, Expression of *STXBP6* along pseudotime (j) and violin/box plots comparing expression in human (h.s.) versus mouse (m.m.) (k). Boxplot center line: Median; Box limits: Upper/lower quartiles.

To evaluate the contribution to distinguish species and find the modules with most contribution, we leveraged the Euclidian and Manhattan distance in the original high dimension feature space. We found that Module 3 and module 9 has the largest distance (Fig. 5d). Permutation test of shows p=0 (Fig. 5e, Methods). We further measured scores of each module along the trajectory the score directly in murine and human (Extended Data Fig. 6a-b). We found several modules as human signature (Module 3, 15) and mouse signature (Module 4, 7). The module score of each sub-cluster derived from mouse and human validate the correctness of its belongings using Seurat. For module3, it shows a strong decline along the trajectory (Pseudo-time Bin) (R^2 = 0.848, p < 0.001) (Fig. 5f), indicating the relationship with human specific signature. Function annotation is further analyzed (Fig. S5) and Module 3 enriched in axon and synapse related function. We further investigated the morans I index which indicate the spatial variation for each gene along the trajectory. There is a great mount of module3 genes ranking at the top with highest morans I. In addition, we found that the overall DS and morans_I was positively correlated with each other (Fig. 5h). We found *NTNG1*/*Ntng1* (Netrin-G1) has been shown to regulate fear-like and anxiety-like behaviors in dissociable neural circuits (ref^49^). we profile that the *NTNG1*/*Ntng1* display a high divergence among the two species (Fig. 5h-k), with higher expression in human, indicating the value of this gene in the study of PTSD in human model.

### Trajectory Analysis of L2/3IT Neuron Reveals Modules Related with Evolution from IL PFC to vmPFC

To investigate cross-species transcriptional trajectories in the IL PFC and vmPFC, we again focused on excitatory L2/3 neurons and examined their variation from mouse to human (Fig. 6a). Using the same analytical pipeline, we deliberately selected pseudotemporal paths that reflect a gradient of nuclei transitioning from one species to another. In the IL PFC to vmPFC comparison, we identified a trajectory along which the normalized proportion of human nuclei increased significantly (R² = 0.741, p < 0.001), accompanied by systematic gene expression changes along this species-specific continuum.

**Figure 6.**
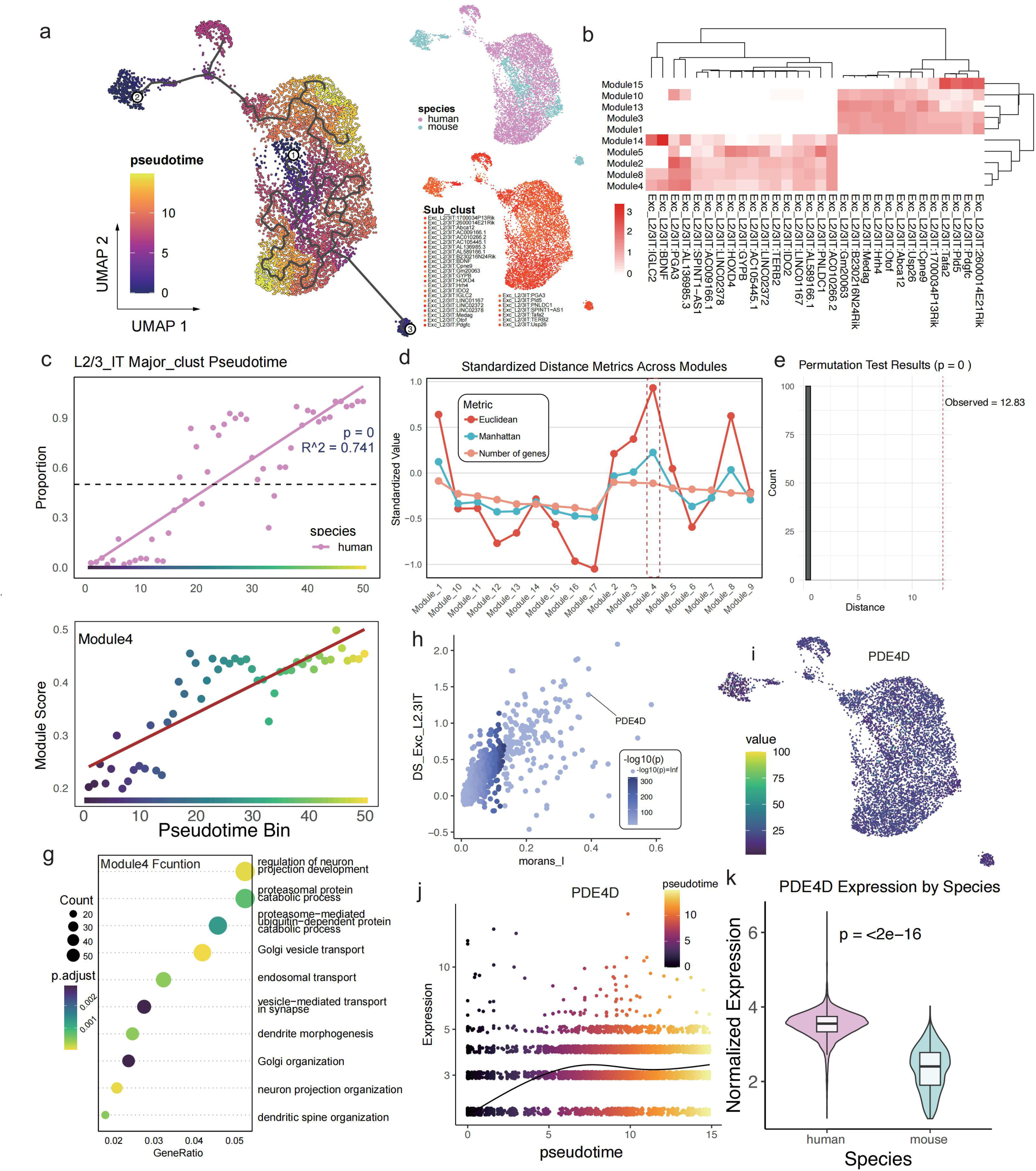
Trajectory analysis of L2/3IT excitatory neurons from mouse IL to human vmPFC. a, UMAP of integrated mouse and human L2/3IT neurons, colored by pseudotime inferred from defined Start Points 1 & 2. Top right: Species origin. Bottom right: L2/3IT subclusters. b, Module activity scores from NMF of trajectory DEGs across subclusters. c, Proportion of human cells within pseudotime bins. Red line: Linear fit. d. Change in scaled Euclidean/Manhattan distances upon module removal. Red dashed box highlights module 4. e, Permutation test p-value (p=0) for module 4 removal effect. f, Module 4 activity score across pseudotime bins. Red line: Linear fit. g, Bubble plot of GO enrichment for genes within module 4. h, Scatter plot of Moran’s I versus DS for trajectory DEGs. i, Feature UMAP plot showing expression of *LDB2* (a module 4 gene) in L2/3IT neurons. j-k, Expression of *LDB2* along pseudotime (j) and in human vs mouse (k). Format as in Fig 5j-k.

By profiling module scores across sub-clusters and directly comparing the average module scores between murine and human cells, we identified modules 8, 14, 4, 2, and 5 as enriched in human nuclei, while modules 10, 15, 13, 3, and 1 were enriched in mouse nuclei (Fig. 6b; Extended Data Fig. 7a-b). Compared to the trajectory between PL PFC and dACC, the IL PFC-vmPFC trajectory exhibited increased species resolution, as evidenced by clearer UMAP separation (Fig. 6a) and higher R² values (Fig. 6c).

Furthermore, by quantifying the contribution of each module to species separation, we found that module 4 displayed the highest discriminative power, with a permutation test yielding p = 0 (Fig. 6d–e). Functional enrichment analysis revealed that module 4 was significantly associated with neuronal projection development and vesicle-mediated transport (Fig. 6g; Extended Data Fig. 7c). Notably, module 4 includes *PDE4D/Pde4d* and *HTR2C/Htr2c*, both of which exhibited high Moran’s I values and have previously been implicated in PTSD and anxiety-related behaviors in mouse models (refs^50,51^). These findings suggest a translational potential for these genes in understanding human PTSD vulnerability.

### Cell Type Contribution in Human PFC to PTSD revealed by GWAS Annotated SNP

To integrate our human multi-omic data with disease risk, we leveraged PTSD GWAS summary statistics (ref^52^). Because genes that harbour many SNPs with low GWAS p-values might be more strongly associated with disease, we speculated that a cell type whose robust marker genes or marker chromatin peaks overlap such disease-linked loci is likely to contribute to pathogenesis. Using MAGMA (ref^53^) and East-Asian reference data from the 1000 Genomes Project (ref^54,55^), we annotated SNPs to genes and quantified, for each brain region, the probability that individual cell types influence PTSD (Methods).

Among glial populations, oligodendrocyte precursor cells (OPCs) from both the dACC and vmPFC exhibited a significant association with PTSD. Within inhibitory neurons, *PVALB* interneurons in both regions, *NDNF* interneurons in the dACC, and *VIP* interneurons in the vmPFC were likewise implicated. For excitatory neurons, vmPFC L6CT, dACC L5ET and vmPFC L2/3IT subtypes contributed significantly to PTSD risk (Fig. 7a).

**Figure 7.**
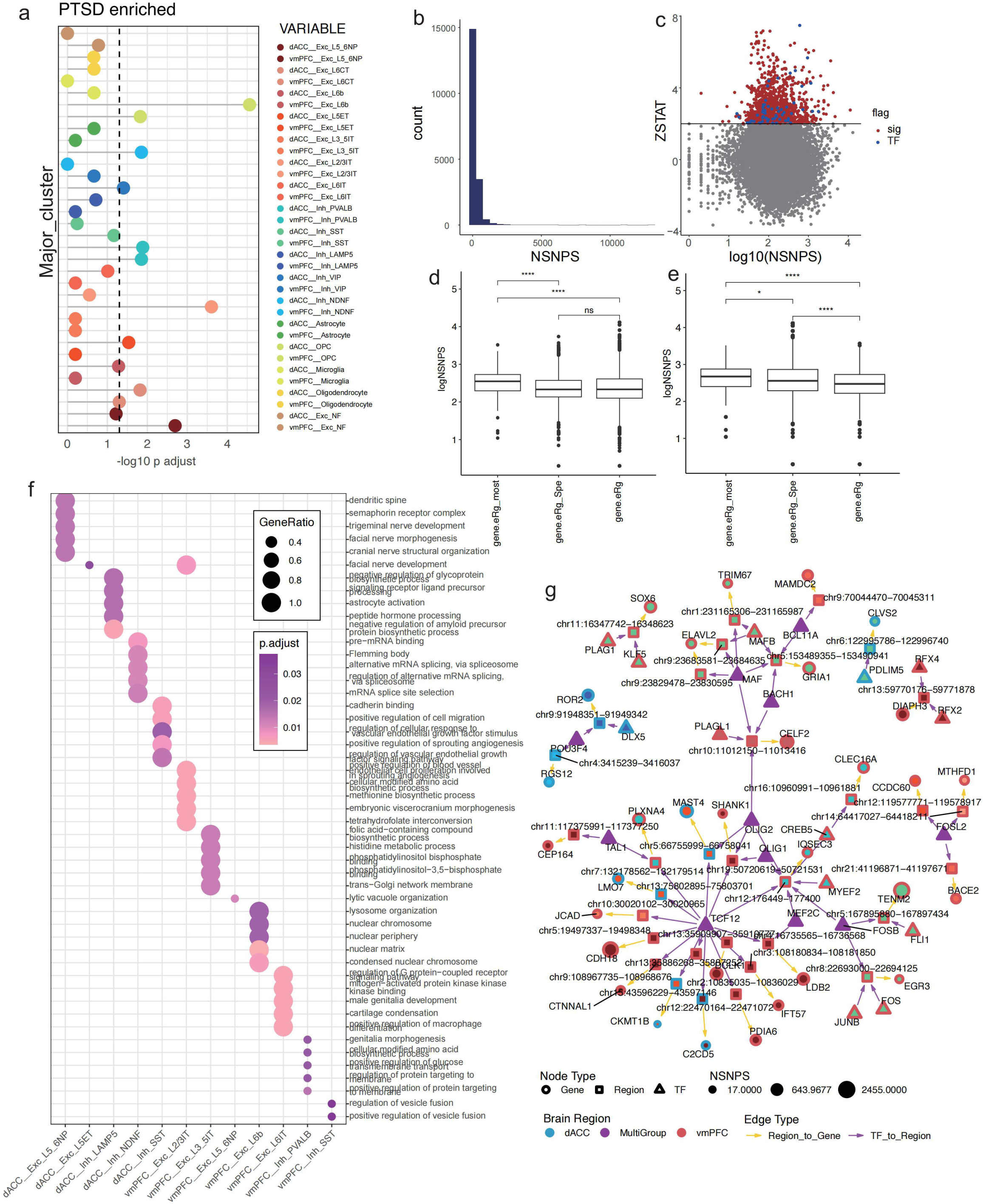
PTSD-associated regulatory networks in human vmPFC and dACC. a, MAGMA analysis assessing the contribution of cell-type-specific subregional (dACC/vmPFC) marker genes and peaks to PTSD risk using East Asian GWAS summary statistics. X-axis: -log10(adjusted p-value). b, Histogram of the number of SNPs within cell-type-specific subregional marker genes and peaks across all cell types. c, Scatter plot of SNP count versus ZSTAT (a measure of association strength; ZSTAT > 2 indicates significance) for PTSD-associated loci. Red: Significant genes; Blue: TFs. d-e, Number of SNPs within genes regulated by eRGs of varying specificity levels (eRg, eRg.Spe, eRg.Spe.Most) in dACC (d) and vmPFC (e). Significance: * p < 0.05; **** p < 0.00005. f, Bubble plot of GO enrichment for genes significantly associated with PTSD risk within each cell type. Format as in Fig. 1j. g, Potential PTSD-associated regulatory network. Nodes: Triangles (TFs), Squares (Regulatory regions), Circles (Target genes). Color: Cell type origin of the eRG. Purple TFs: Function in multiple cell types. Stroke color: Subregion origin (dACC/vmPFC). Circle size: Number of GWAS SNPs within the gene.

Most genes carried fewer than 500 PTSD-linked SNPs (Fig. 7b). Genes surpassing the significance threshold (ZSTAT > 2) were markedly enriched for transcription factors. When we stratified eRg by cell-type specificity, the numbers of SNPs mapped to both regions and genes rose markedly with increasing specificity, indicating a positive relationship between SNP burden and highly specialised eRg.

Functional annotation of marker features that overlapped significant GWAS genes revealed distinct pathways for each dACC and vmPFC cell type (Fig. 7f). Gene-regulatory networks built from GWAS-significant genes highlighted TCF12 as a central hub in both neuronal and glial modules (Fig. 7g). Notably, factors such as *MEF2* and *FOSB*—previously linked to neuropsychiatric phenotypes and fear-related behaviours (ref^56,57^)—also occupied prominent positions. The preponderance of genes, regulatory regions and transcription factors originating from the vmPFC suggests that loss-of-function variants may accumulate more frequently in this region than in the dACC. Meanwhile, we also found a complex regulation network for glia cells, suggesting the potential function of glia in the regulation of PTSD (Extended Data Fig. 8a-b).

### Chromatin Primed Genes Reveals the Suspicious Genes Related with PTSD

Chromatin primed genes (CPGs) are defined as genes with high accessibility but remain low expressed at RNA level, plays a vital role in neuron development and diseases (ref^58^). We hypothesis that CPGs, under certain stress, could be activated and start transcription. To find the CPGs which might contribute to PTSD, we leveraged the data from vmPFC PTSD patient bulk RNA-seq and found the genes that remain silent in naïve donors but actively transcribed in PTSD patients’ vmPFC (ref^8^). We discovered that the overall gene expression and eRg region’s accessibility is overall positive related with each other. When using the regions and genes defined in eRg across all cell types and eRgs, there is an overall positive correlation: as accessibility increases, so does expression. However, a closer examination discloses that within the low-accessibility phase—from bin□1 to bin□2—an increase in enhancer-region accessibility does not lead to a corresponding rise in expression. Instead, expression decreases, most notably in L6CT and L2/3IT excitatory neurons. This pattern is not observed in the dACC region (Fig. 8a). When directly measuring the gene expression and chromatin accessibility of the gene itself, the positive regulation is more positive related with each other (Fig. 8b). For each cell type, the data reveal a strict positive correlation between the activity score and the expression score. Moreover, during the transition from bin□4 to bin□5, there is a pronounced increase in both metrics.

**Figure 8.**
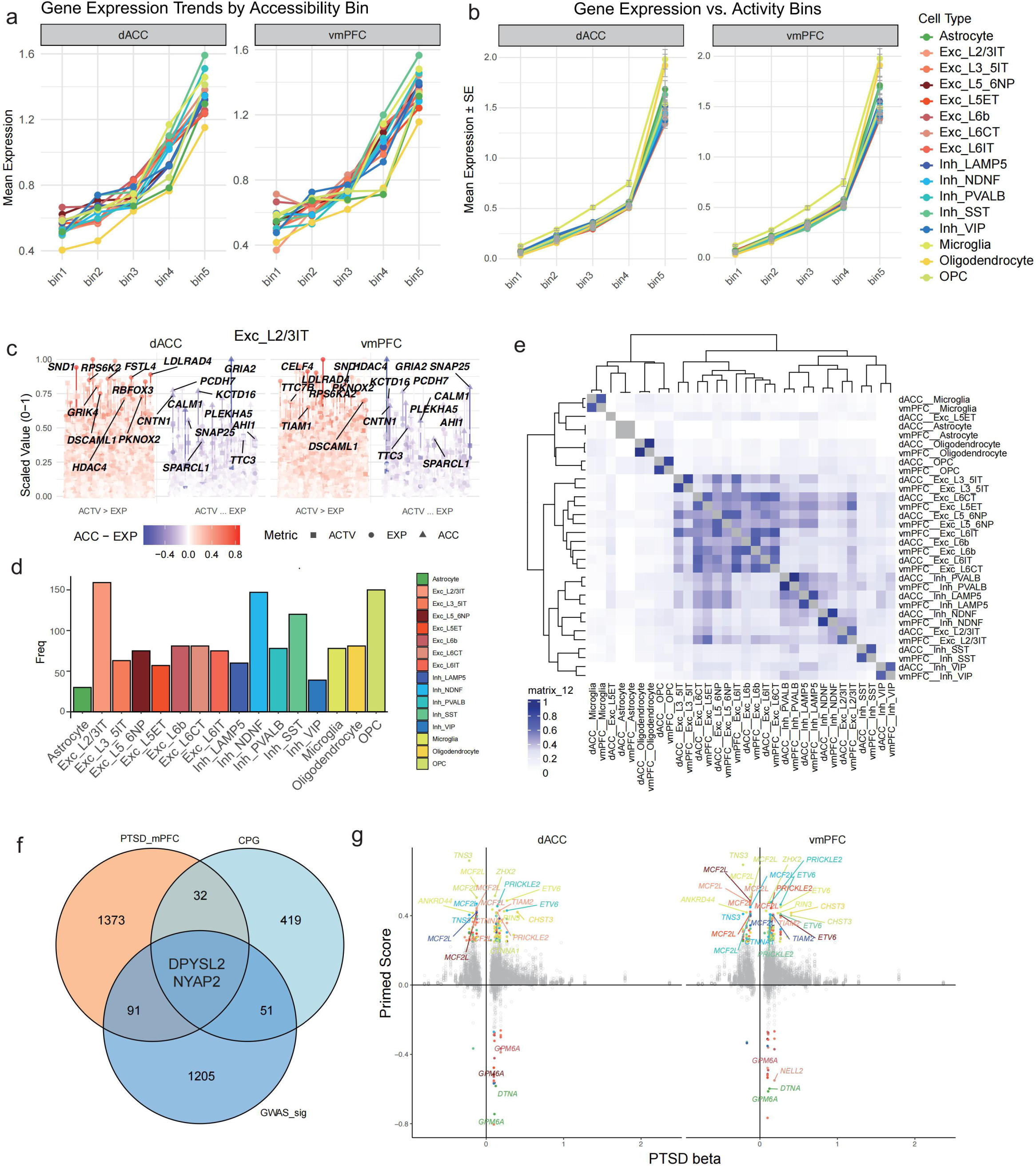
Chromatin Primed Genes in human PTSD. a. Gene expression value and the accessibility of TF binding regions (bin1∼5) in dACC and vmPFC. b. Gene expression value and the accessibility of gene (bin1∼5) in dACC and vmPFC. c. The profiling of gene’s accessibility, TF binding region accessibility and gene expression (all scaled). Each vertical column represents three attributes of a single gene: overall chromatin accessibility (ACTV, circles), gene expression level (EXP, triangles), and enhancer accessibility (ACC, squares). For every cell type, the values of these three attributes are min–max scaled to a 0–1 range. Genes are divided into two groups according to the relative values of ACTV and EXP: genes with ACTV > EXP are placed in group 1, whereas genes with ACTV < EXP are placed in group 2. Consequently, genes in group 1 are considered candidate chromatin-primed genes (CPGs). Columns are colour-coded by the scaled difference (ACTV − EXP); deeper red indicates a higher degree of chromatin priming. d. Barplot of the number of unique CPGs in each major cell type. e. Similarity measured by Jaccard Index of the CPGs in each major cell type. f. VennDiagram of the intersection of GWAS significant genes, CPGs and PTSD up-regulated genes found by PTSD patients vmPFC bulk-RNA-seq. g. Integrated analysis of CPGs and PTSD patients vmPFC bulk-RNA-seq, x-axis represents ths beta value (derived from fold-change) while y-axis represents the PrimedScore.

We further measured the chromatin prime level in each cluster and brain region. Each vertical column represents three features of a given gene: overall chromatin accessibility (ACTV), depicted as a circle; gene expression level (EXP), shown as a triangle; and enhancer region accessibility (ACC), represented by a square. The values of these three features are scaled from 0 to 1 within each cell type. Genes are classified into two groups based on the relationship between ACTV and EXP: if ACTV is higher than EXP, the gene is assigned to Group 1; if ACTV is lower than EXP, it is assigned to Group 2. Genes in Group 1 are considered candidate primed genes (CPGs). The color of each gene marker indicates the difference between scaled ACTV and EXP values, with deeper red tones representing a higher degree of “priming” (Extended Data Fig. 9). We highlight the primed genes identified in a L2/3IT excitatory neuron. These genes show a relatively high degree of consistency between the dACC and vmPFC, suggesting conserved regulatory priming across these brain regions (Fig. 8c). We found in total 504 unique CPGs from each brain region and cell type (PrimedScore: ACTV-EXP > 0.25 & EXP < 0.25) (Fig. 8d). The overall similarity measured by JI reveals that the candidate primed genes (CPGs) show high similarity between the two brain regions. Meanwhile, there is minimal overlap in CPGs between glial cells and neurons. Also, the greatest CPG similarity is observed within excitatory neuronal subtypes. These findings suggest that CPG classification is strongly associated with the biological identity of the cell. In other words, cells sharing the same biotype tend to exhibit similar sets of primed genes.

Function annotation of the CPGs shows that there are consistent terms for each cluster while still unique function for CPGs for each cell type (Extended Data Fig. 1a). The functions of CPGs mainly fall into synapse or dendrite related function for neurons and Glia, suggesting that CPGs in glia that contribute to PTSD is related with neuron function. All these Indicate the potential neuron-glia function in the pathogenesis of PTSD.

We also found that from PTSD GWAS gene with high SNPs, CPGs and DEGs from PTSD patient vmPFC region gives 2 genes, including DPYSL2, which is a microtubule□stabilizing protein essential for neuronal development (ref^59^), and NYAP2 which is essential in neuron projection morphogenesis(ref^60^) (Fig. 8f). While integrating CPGs with DEGs from PTSD patient, we found that in each brain region, there is a substantial number of PTSD DEGs exhibit higher chromatin accessibility than expression levels (PrimedScore□>□0), indicating various degrees of priming among PTSD-associated genes. We can also pinpoint in which specific cell types this priming occurs. Additionally, there are some PTSD DEGs that are actively expressed (i.e., with detectable transcription). There are several genes that are primed in naïve and up-regulated in patient such as TIAM2, which regulates dendritic spine and growth cone formation, neurite outgrowth (ref^61,62^). Function annotation of genes “primed in naïve and up-regulated in patient” gives us close relationship to actin-based cell projection, cell migration and glia guided cell migration (Extended Data Fig. 11b), while those consistently highly expressed genes are enriched in functions including growth cone and axon related functions (Extended Data Fig. 10c).

## Discussion

In this research, we enrolled brain regions with close relationship with PTSD-PLPFC/ILPFC in mouse and dACC/vmPFC in human. We compared the difference within species with respect to the gene expressing and chromatin accessibility, combining which we found the cell-type specific gene regulation network and revealed the regulation patterns that differ in regions and species. We further compared the similarity and difference for homolog cell-types utilizing all features, marker features and ortholog genes with divergent score. We found the evolution modules that contribute most to the cell transforming from mouse to human using excitatory layer 2/3 IT neuron in each brain region, respectively. Finally, using PTSD GWAS data and vmPFC bulk-RNA-seq data, we successfully found cell types with high contribution to PTSD with respect to SNPs and chromatin primed genes that are highly expressed under stress.

In our human-derived data, the number of glias increased significantly comparing that in mouse. While Recent research has challenged long-held beliefs about the proportion of glial cells to neurons in the human brain (Contrary to the widely cited 10:1 glia-to-neuron ratio, studies using advanced counting methods like the isotropic fractionator have revealed a ratio closer to 1:1) (ref^63,64^). However, there still has been argues about the oligodendrocyte number along aging. There has been study shown that number of oligodendrocytes increases by about 50% in the aging primate cerebral cortex, likely due to generation from progenitor cells (ref^65^). Meanwhile, there is also studies showing that oligodendrocyte numbers decrease with age (ref^66^) and in very old individuals (ref^67^). In our research, doners were above 60 years old, with a higher proportion of oligodendrocytes comparing with the researched enrolled younger individuals (ref^68^).

Excitatory neurons marked by neurofilament expression exhibit high levels of canonical excitatory neuronal markers, while also displaying transcriptional signatures that suggest a close association with oligodendrocytes. Although we did not investigate the developmental origin or specific functional roles of this cluster, it represents a distinct population warranting further characterization. Future studies could provide deeper insights into the biological significance and potential functional specialization of this hybrid-like cell type. Additionally, we observed that mitochondrial genes may be retained during single-nucleus sequencing due to their perinuclear localization. Rather than being discarded as technical artifacts, these mitochondrial transcripts could potentially serve as indicators of cellular states or metabolic activity.

Trajectory analysis is widely applied across various single-cell and single-nucleus studies, despite the fact that only a minority of these studies incorporate true time-series data. In this work, we demonstrate how Monocle3 can be effectively utilized in non-time-series datasets by strategically selecting start point(s) such that the distribution of cells or nuclei along the resulting pseudotime trajectory reflects a meaningful gradient associated with experimental variables. This approach enables the abstract concept of “pseudotime” to be anchored to biologically relevant differences, thereby enhancing its interpretability and facilitating downstream analyses guided by trajectory-based insights.

Despite the single-cell atlas of human and mouse prefrontal cortex and the investivation of multi-omics in PTSD research (ref), few research concerns about the two brain regions in prefrontal cortex which has diverse function. Our research bridges the gap of understanding the cell-type and gene regulation difference in the 2 brain regions, and further more, using correspondent brain region, we build the paralle human sub-region atlas, in order to reveal the gene regulationnsimilarity along with the divergent from mouse to human, thus provide reference for researchers to find key molecules that has the potential transfering value to human-related research.

In our single-cell transcriptomic data, inhibitory neurons were not only markedly fewer in number compared to excitatory neurons, but also exhibited higher transcriptomic similarity between the PL and IL regions. As a result, fewer transcriptionally distinct inhibitory neuron subtypes were identified relative to excitatory neurons. In contrast to the PL, the IL region showed a relatively higher abundance of astrocytes and oligodendrocytes. Moreover, glial activation trajectory analysis revealed that glial cells in the IL were more transcriptionally active in functions related to cellular structure and morphology maintenance. L2/3 IT neurons, the most abundant and diverse subtype in both PL and IL, show complex connectivity patterns and region-specific functions. Trajectory and regulatory network analyses revealed a shift from synaptic signaling in PL to synaptic structural maintenance in IL. Spatial transcriptomic analyses further supported this functional divergence, with PL signatures enriched for synaptic signaling and IL signatures linked to structural maintenance and glial activity. Together, these findings suggest a potential mechanism by which IL contributes more prominently to memory retention, particularly in the context of fear extinction.

The regulation network is the graph from the aspect of mathematics. There have been multiple researches trying to reveal the different regulation network inside of it (ref^69^). In this study, rather than focusing on identifying distinct regulatory networks, we aimed to uncover shared or common regulatory networks across brain regions, defined by regulatory elements that can be bound by multiple transcription factors and regulate multiple genes. We constructed region-based network components by integrating marker genes and marker regions derived from snRNA-seq and snATAC-seq data, respectively, to annotate eRegulons. This approach offers high flexibility, as it relies solely on cluster-specific marker features, thereby enabling the annotation of sub-cluster-level-specific eRegulons. Notably, there is currently no established benchmark for using the SCENIC+-provided feather files, and we observed that leveraging custom-built feather files substantially improved the resolution and quality of the results.

Our research, in summary, provide a comprehensive atlas of human and mouse prefromtal crotex sub-regions, and investigated the cross-species similarity and divergency, revealed the dynamic gene expression, chromatin accessibility and regulation network, and shed light on the contribution of regional cell-types to PTSD along with the chromatin primed genes related with PTSD.

## Methods

### Sample Collection for Mouse Brain Tissue

Male C57BL/6J mice (4 pooled together for each sample), aged 8 weeks at the experiment dat, were housed in standard home cages for two weeks prior to the experiment. All animals were maintained under standard conditions with ad libitum access to food and water, and a regular light-dark cycle. Environmental noise was strictly minimized throughout the housing period, and all experimental procedures were preferentially conducted during the daytime. In accordance with institutional animal facility protocols, mice were transported between housing and sampling rooms using disposable light-shielded white boxes to prevent external stress. Prior to tissue collection, animals were temporarily held in the designated transport area outside the sampling room.

Each mouse was individually brought into the sampling room and immediately sacrificed. The head was severed using scissors, and the skull was carefully removed to extract the whole brain, including the olfactory bulbs, cerebrum, cerebellum, and part of the brainstem. The intact brain was transferred onto a clean glass slide and snap-frozen in a dry ice-cooled container.

A brain slicer (RWD Brain Matrix, #68707) was pre-chilled on ice. The brain was oriented ventral side up and placed into the matrix, with the olfactory bulbs aligned to the designated anterior groove. A double-edged stainless steel razor blade (Claude brand) was inserted from the posterior end to stabilize the brain. The first blade was inserted at a 45° angle into the first anterior slot, cutting vertically through the tissue. A second blade was inserted into the third slot using the same approach, followed by a third blade placed between the first two. The three blades were gently aligned and simultaneously pressed down to ensure complete separation of the targeted brain regions. All three blades were then lifted together in a single motion. The blades were carefully separated to yield two coronal brain slices, each 1 mm thick, encompassing the infralimbic (IL) and prelimbic (PL) regions of the prefrontal cortex (PFC). Dissection tools such as microforceps or glass needles were used to assist with separation if needed.

The slices were transferred to clean, pre-chilled glass slides (note: the slides were not pre-cooled on dry ice), and labeled accordingly. Reference maps of IL and PL regions were prepared in advance using the Allen Brain Atlas (ref^70^). Slides were placed on a chilled metal surface and examined under a dissection microscope. Using fine microsurgical tools, the IL and PL PFC regions were carefully isolated. Tissue samples were immediately placed into labeled 1.5 mL microcentrifuge tubes and snap-frozen on dry ice. This procedure was repeated identically for all mice within the same experimental batch.

Importantly, mice that had not yet been processed were not permitted to enter the sampling room and were instead held in a quiet, dark environment outside the room to avoid stress-induced artifacts.

### Sample Collection for Human Brain Tissue

Postmortem human brain tissues (5 males aged 60-85 y/o were obtained from the department of Anatomy, Huazhong University of Science and Technology. All experiments were approved by the Institutional Review Board of Huazhong University of Science and Technology (Approval No.: 2024-S124) and performed in accordance with the principle of the Helsinki Declaration II. Written informed consent was obtained from each participant.

Upon harvesting, brain tissues were immediately stored at −80□°C. Target brain regions, including the ventromedial prefrontal cortex (vmPFC) and dorsal anterior cingulate cortex (dACC), were sectioned into thick coronal slices. Dissections were performed using a scroll saw under the guidance of a human brain anatomical atlas and supervised by Professor Xingyan Li (HUST), a certified expert in neuroanatomy.

Throughout the entire procedure, samples were maintained in a frozen state. Tissue blocks were further fragmented using a pre-chilled mortar and pestle, ensuring that sample integrity was preserved and thawing was strictly avoided at all stages.

### Single-nucleus RNA and ATAC Library Preparation and Sequencing

Single-cell RNA-seq libraries were constructed using the Chromium Single Cell 3’ Library & Gel Bead Kit v3.1 (10x Genomics, 1000121) and the Chromium Single Cell G Chip Kit (10x Genomics, 1000120). A cell suspension with a concentration of approximately 300–600 viable cells per microliter—quantified using the Countstar system—was loaded into the Chromium Controller (10x Genomics) to generate single-cell gel bead-in-emulsions (GEMs). Each cell was encapsulated in an individual GEM droplet, suspended in PBS containing 0.04% BSA to maintain cell integrity. For each reaction channel, approximately 6,000 cells were loaded, with an expected recovery of around 3,000 individual cells. Following encapsulation, cells were lysed within GEMs to release RNA, which was subsequently reverse transcribed and uniquely barcoded inside each droplet.

Reverse transcription was performed on an S1000™ Touch Thermal Cycler (Bio-Rad) under the following conditions:1. 53□°C for 45 minutes, 2. 85□°C for 5 minutes, 3. hold at 4□°C.

After reverse transcription, barcoded cDNA was recovered from the emulsions and amplified. Quality and fragment size distribution of the amplified cDNA were assessed using the Agilent 4200 TapeStation system prior to library construction.

For snATAC-seq, nuclei were isolated following the protocol “Nuclei Isolation for Single Cell ATAC Sequencing” provided by 10x Genomics (CG000169). Briefly, tissues were rapidly processed and lysed on ice using pre-chilled LB Buffer. Frozen or freshly dissected tissues were minced using scissors or homogenized with a tissue homogenizer in 1 mL of LB Buffer in 2 mL centrifuge tubes. The lysate was incubated on ice for 1–10 minutes, with the lysis duration optimized based on tissue type in pilot experiments. The lysate was then passed through a 40 μm cell strainer to remove debris, and the filtrate was transferred to a new 2 mL tube and centrifuged at 500□g for 5 minutes at 4□°C.

The supernatant was carefully removed, and the pellet was resuspended in 300 μL of LB Buffer. The nuclei suspension was then transferred to a new tube and mixed with 300 μL of RB Buffer. Subsequently, 600 μL of PB1 Buffer was slowly layered to the bottom of the tube using a pipette tip, followed by an additional 600 μL of PB2 Buffer, also added slowly to ensure clear phase separation. Samples were centrifuged at 4,000□g for 20 minutes at 4□°C. Nuclei were enriched at the interface between the PB1 and PB2 layers and were carefully collected (∼150–300□μL) into a fresh 1.5 mL tube.

To further purify the nuclei, 1 mL of RB Buffer was added, mixed gently, and passed through a 40 μm pipette-tip strainer. The sample was centrifuged again at 500□g for 5 minutes at 4□°C. The pellet was gently resuspended in 100 μL of EB Buffer. If no visible pellet was observed, 100 μL of supernatant was retained to avoid loss of material. Nuclei concentration was measured using a Countstar Rigel S2 automated cell counter. In parallel, 5 μL of the sample was stained with Trypan Blue and observed under a microscope to assess nuclear integrity. Nuclei were maintained in a cold environment throughout the entire process to preserve integrity and prevent RNA degradation.

All buffers (LB, RB, PB1, PB2, EB) were prepared as instructed, and RNase inhibitors (1□U/μL) and DTT (1□mM) were freshly added to all solutions immediately before use. BSA was also added freshly to LB and RB Buffers at a final concentration of 1%. All tubes used were RNase-free (1.5□mL and 2□mL), and all steps were performed on ice unless otherwise stated. For the EB buffer step in the ATAC-seq workflow, the buffer was replaced with the Nuclei Buffer supplied in the Chromium Single Cell ATAC Library Kit.

Immediately following nuclei isolation, single-cell ATAC-seq library construction was initiated according to the manufacturer’s instructions using the 10x Genomics Chromium Single Cell ATAC platform.

For single-cell RNA sequencing, we followed the protocol provided by 10x Genomics and utilized the Chromium Single Cell 3’ Library and Gel Bead Kit v3.1. Libraries were sequenced on an Illumina NovaSeq 6000 platform using paired-end 150 bp (PE150) sequencing. A minimum of 100,000 reads per cell was targeted to ensure sufficient transcriptome coverage.

### Data Pre-processing and Quality Control

Raw sequencing data were processed using the Cell Ranger software (RRID:SCR_017344 10x Genomics), which was obtained from the official 10x Genomics website (https://support.10xgenomics.com/single-cell-gene-expression/software/downloads/latest). The cellranger count pipeline was used to perform alignment, filtering, barcode counting, and UMI quantification, resulting in the generation of feature-barcode matrices and initial cell clustering. Cell Ranger also provided output files for downstream custom analyses, including the sparse expression matrix, barcode files, and gene annotation files.

For downstream analysis, we used Seurat to load each sample individually. An initial round of quality control was performed, in which cells with fewer than 200 detected genes were excluded. We calculated the proportion of mitochondrial gene expression per cell, which was later used for further filtering. All samples were subsequently integrated, and batch effects were corrected. The final merged object included single-cell data from 10 independent batches.

Normalization was carried out using the SCTransform pipeline in Seurat, which models gene expression variance more accurately and improves normalization by accounting for sequencing depth and technical noise. SCTransform returns centered residuals that are used for dimensionality reduction and clustering. However, since gene expression values are transformed through a modeling process, SCTransform-normalized data are not optimal for analyses requiring actual expression levels. For expression-level analyses, we used traditional log-normalization approaches based on sequencing depth. In SCTransform, we applied the glmGamPoi method for model fitting and excluded mitochondrial genes during normalization.

Highly variable genes (HVGs) were identified by assessing gene-wise expression variability across cells, and the top 3,000 HVGs were selected to accelerate downstream analyses such as PCA. To ensure robust downstream gene expression analysis, the raw expression matrix was separately normalized using depth-adjusted log-transformation. SCTransform-normalized data were primarily used for unsupervised clustering and dimensionality reduction. Notably, SCTransform was applied on a per-sample basis before integration to maximize normalization accuracy.

To identify potential doublets—i.e., artifacts from droplets containing more than one nucleus—we employed DoubletFinder (v3) (ref^71^). DoubletFinder simulates artificial doublets by merging expression profiles from two distinct clusters and scores each cell based on its similarity to simulated doublets. Since this method relies on initial clustering information, doublet detection was performed after initial PCA and graph-based clustering (see Section 2.5.4). For clustering, we applied the shared nearest neighbor (SNN) algorithm on SCTransform-normalized data, using a range of resolution parameters (in increments of 0.2). Cluster consistency across resolutions was visualized to determine an optimal resolution. Specifically, we selected the resolution at which subclusters from a parent cluster first split and subsequently merged with unrelated clusters at higher resolutions. This criterion was consistently used for all subsequent cluster identification steps.

Following doublet detection, cells identified as doublets were removed. As batch effects may artificially separate biologically similar cells across different samples, we applied Harmony to further correct for batch variation. Harmony integration was performed on SCTransform-normalized data. Because SCTransform must be applied independently to each dataset before merging, and doublet removal requires prior clustering, the analysis pipeline involved multiple normalization rounds. We summarize the overall workflow below: 1) Load and perform initial quality control on all samples. 2) Apply SCTransform normalization to each sample independently. 3) Merge samples using Seurat’s merge() function (note: not integrate() at this step). 4) Correct for batch effects using Harmony on the merged object. 5)

Identify HVGs, compute mitochondrial gene proportions, and perform SNN clustering using the resolution selection criteria described above. 6) Apply DoubletFinder to remove doublets based on initial clustering. 7) Split the doublet-filtered object by sample and repeat steps 2–5 to obtain the final integrated dataset.

Dimensionality reduction was performed using PCA on SCTransform-normalized data following the pipeline of hbctraining (https://zenodo.org/records/4783481). To select the number of significant PCs, we plotted the variance explained by each PC (elbow plot). The number of retained PCs was determined by taking the larger value from the following two criteria: 1) The first PC for which the proportion of variance explained dropped below 5% and the cumulative variance exceeded 90%. 2) The first instance in which the difference in variance explained between two consecutive PCs fell below 0.1%.

The selected PCs were then used for nonlinear dimensionality reduction via UMAP and for downstream clustering.

### Cell-type Annotation and Sub-type Identification

#### Cell-type Annotation

Cell type annotation was performed by mapping the unsupervised clusters generated through the shared nearest neighbor (SNN) algorithm to canonical cell types. Since the number of clusters identified by unsupervised algorithms often exceeds the number of known biological cell types, we first identified marker genes for each cluster and refined these markers using the starTracer framework.

Cell type assignment was then determined through the integration of the following strategies:1) Marker-based comparison: Optimized cluster-specific marker genes were systematically compared to curated marker gene sets for known cell types to establish correspondence. 2) Expression-based visualization: Canonical marker genes for major cell types were visualized across clusters to infer the most probable cell type identity for each cluster. 3) Reference-based mapping: The Azimuth platformwas employed to project single-cell profiles onto a reference atlas of the mouse cortex, enabling high-resolution cell type predictions.

By integrating these three approaches, we achieved robust and biologically consistent cell type annotations at two hierarchical levels: 1) Broad biological classes (Bio_clsust), including excitatory neurons, inhibitory neurons, and non-neuronal cells. 2) Fine-grained cell types (Major_clust), encompassing seven excitatory neuronal subtypes, five inhibitory neuronal subtypes, and four types of glial cells as well as endothelial cells.

For mouse and human homolog cell types, we utilized RPCA from Seurat to transfer labels using the annotaed mouse data, there has been a slightly difference in the combination of inhibitory neuron marker gene and layer 4 excitatory neurons. Exc_L3_5IT neuron will corresponds to mouse Exc_L3/5IT neuron, while inhibitory *Sncg* neuron in mouse corresponds to *NDNF* neuron in human.

Marker gene selection was guided by previously published single-cell transcriptomic studies of both mouse and human cerebral cortex, as well as canonical marker genes recorded in the Azimuth reference database. For additional marker discovery, we applied Seurat’s default differential expression framework. Specifically, for each cell type or subcluster, all remaining cells were grouped into a single reference population, and differential expression was assessed using the Wilcoxon rank-sum test.

Only upregulated genes were considered, and candidate markers were filtered using an adjusted p-value threshold of 0.05 and a minimum average log□ fold change (avg_log2FC) greater than 0.25. The resulting gene lists were further refined using the starTracer (ref^26^). From the optimized output, the top five ranked genes were selected as representative markers for each cell type. For visualization, violin plots were generated using the top three marker genes per cluster.

#### Sub-type Annotaiton

In previous studies, the choice of K (i.e., the number of clusters) has often relied on subjective estimation, where researchers visually assessed how many “groups” appeared to be present in the data. To systematically determine the optimal K values for both snRNA-seq and snATAC-seq datasets, we employed MultiK (ref^32^), a data-driven tool for robust K selection. For each annotated major cell type, the corresponding subset of the snRNA-seq and snATAC-seq objects was extracted and imported into MultiK for analysis.

To account for the notion that an optimal clustering solution should be stable against small perturbations, we performed random subsampling of each dataset, retaining 80% of the original cells. Clustering was conducted using a shared nearest neighbor (SNN) graph-based approach, with resolution parameters ranging from 0.05 to 2.0 in increments of 0.05. For each resolution, multiple clustering runs were performed, and the relative principal average connectivity (rPAC) scores were calculated.

MultiK then inferred the optimal K by analyzing the frequency distribution of observed K values against their corresponding (1 – rPAC) scores. Scatter plots were generated to visualize the relationship between clustering robustness and K frequency, thereby identifying the most stable and consistent clustering solutions across subsampled iterations.

### Sptial Transcriomics, sample preparation and data analysis

Brain tissue collection was partially conducted following the procedures described before. Prior to dissection, mice were anesthetized using pentobarbital. Following anesthesia, mice were perfused with 1% paraformaldehyde (PFA). After complete perfusion, brain tissues were harvested at room temperature. Notably, for spatial transcriptomics, the brains were not sectioned using a brain mold. After collection, tissues were subjected to a graded ethanol dehydration process.

After switching on the main power and completing system self-check, the embedding station was configured according to experimental needs. Parameters such as paraffin reservoir temperature, cassette holding chamber temperature, module heater temperature, working surface temperature, working dates, operation start/stop times, and selected working days were set in the configuration mode. Once set, the instrument retains the selected parameters unless manually changed.

When all components reached the preset temperatures and the paraffin was fully melted, embedding began. Dehydrated tissues in their cassettes were transferred from the dehydrator to the cassette storage chamber. Organs from the same animal were placed on the preheated working surface with a small amount of paraffin. Embedding molds were positioned under the paraffin dispenser, and paraffin embedding was performed using the foot pedal control. Upon completion, the workstation was cleaned and cleared of debris.

The workstation must be stable and level. A rear spacer bar must be installed to ensure a minimum 15 cm distance between the instrument and the wall, and no air conditioning vents should be present nearby. Newly added paraffin should not exceed 70°C, as excessive heat may damage critical instrument components and compromise embedding quality.

As paraffin is flammable, care must be taken to avoid spills. Any leaked or residual paraffin should be removed using a designated scraper, not sharp instruments, to prevent damage to the instrument surface. The main power switch should not be frequently toggled; daily operations should be controlled via the interface panel. The main switch should only be turned off when the system is not in use for extended periods.

Embedded brain tissues were sectioned using a microtome. Each section was examined based on anatomical coordinates from the Allen Brain Atlas. Appropriate sections were carefully selected, lifted using a fine brush, and floated on the surface of water to allow natural expansion. Once fully expanded, 10x Genomics Visium Spatial Gene Expression slides were immersed in the water, enabling the tissue section to attach to the slide surface.

Spatial transcriptomics samples were processed using Seurat. Each sample was independently subjected to SCTransform normalization, following the procedures described in Sections 2.5.1 to 2.5.4, with the exception that doublet removal was not performed. Following normalization, spatial transcriptomics data were mapped onto the corresponding single-cell transcriptomic reference using Seurat’s built-in anchoring framework for single-cell to spatial data integration. This enabled the assignment of spatial spots to distinct cell clusters with improved accuracy.

Due to the current resolution limitations of spatial transcriptomics platforms, each spatial spot typically captures transcripts from tens of cells, resulting in what is effectively a miniature bulk RNA-seq profile. However, with access to a matched single-cell RNA-seq reference from the same tissue region, the gene expression profile of each spot can be approximated as a linear combination of the expression profiles of its constituent cell types. This allows for a computational decoding of each spot’s cellular composition—a process known as deconvolution.

To achieve this, we applied CARD (ref^72^), a widely used deconvolution method for spatial transcriptomic data, leveraging the single-cell gene expression reference to infer the proportional contributions of each cell type within each spot. Additionally, a Bayesian-based deconvolution approach was applied to further refine gene expression estimates, resulting in improved resolution of both cellular composition and transcript abundance across spatial coordinates.

### Diversity Across Brain Regions

To evaluate the regional distribution differences of each cell subcluster between IL PFC and PL PFC, the integrated Seurat object was first divided into two subsets based on the sample origin (IL PFC or PL PFC). For each annotated cell subcluster, we counted the number of cells in both IL and PL PFC. To correct for differences in sequencing depth and total cell recovery between regions, we normalized the cell counts by the total number of cells within each respective cortical region. The relative abundance ratio between PL and IL was then calculated as:

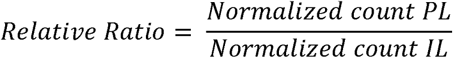

A ratio close to 1 indicated a comparable distribution between IL and PL PFC, whereas a value >1 suggested an enrichment in PL PFC, and <1 indicated enrichment in IL PFC.

To spatially visualize these regional differences on the UMAP embedding, we extracted the UMAP coordinates of cells from IL and PL separately. A 1000 × 1000 grid was defined to overlay the UMAP space. The three key points — (0, 0), (1000, 0), and (0, 1000) — were linearly mapped to the minimum point, maximum UMAP x-axis value, and maximum y-axis value, respectively, thereby aligning the grid to the coordinate space of UMAP embeddings.

We then used the MASS package to calculate the cell density within each grid cell for IL and PL PFC independently. By subtracting the IL density map from the PL density map, we obtained a Relative Distribution Score (RDS) for each grid cell. This score reflected the relative spatial enrichment of cells between PL and IL.

Next, the UMAP coordinates of all cells were again mapped to the corresponding 1000 × 1000 grid, and based on their location, each cell was assigned the RDS of its grid cell. This annotated score was written back to the metadata of the Seurat object using the cell barcode. The resulting annotation enabled us to visualize regional distribution changes directly on the UMAP plot, highlighting spatial shifts in cell population localization between PL and IL PFC.

To investigate transcriptional changes across different behavioral phases, we used Naïve samples as the control group, and grouped all cells according to their tissue origin and behavioral stage (e.g., Naïve, Fear Conditioning [FC], and Extinction [EXT]). For each annotated cell subcluster, we performed differential expression gene (DEG) analysis using the Wilcoxon rank-sum test, comparing the transcriptomes from the Naïve state to FC and EXT conditions respectively.

To comprehensively capture all candidate differentially expressed genes (DEGs) and differentially accessible regions (DARs), we initially set the p-value threshold to p = 1, allowing the inclusion of all genes for subsequent filtering and visualization. We then applied a uniform filtering threshold across all subclusters, selecting significantly regulated genes using the following criteria: adjusted p-value < 0.05, and average log2 fold change > 0.25.

The number of upregulated DEGs was then quantified for each subcluster to assess the extent of transcriptional activation during memory formation and extinction.

### Identification of eRegulons

To reconstruct gene regulatory networks from our multiomic single-cell data, we utilized the SCENIC+ pipeline (ref^39^)—an advanced framework that integrates single-nucleus RNA and ATAC sequencing to infer enhancer-driven regulatory programs. We began by exporting our processed Signac object from snATAC-seq analysis and preparing it via pycisTopic for chromatin topic modeling. snRNA-seq data was processed using scanpy (ref). Differential accessible regions were identified using criteria of |log□FC| > 0.25 and adjusted p < 0.05, including both cell-type-specific marker regions and global differential peaks.

Following SCENIC+ practices, we generated consensus peak- and motif-based feather files to prepare inputs for motif enrichment and region-to-gene linking. Both direct and extended eRegulon definitions were included in downstream analysis.

Cell-type specificity of each eRegulon was determined by requiring that both the transcription factor’s binding region and its predicted target gene annotation originated from the same cell cluster. This yielded cell-type-specific eRegulons. To prioritize the most discriminating regulators, we quantified each eRegulon’s specificity using an entropy-based metric: information entropy, adapted from information theory, which could be used in assessing regulon specificity.

Finally, we computed a triple_rank score—a composite metric that integrates three elements: TF motif enrichment, region accessibility, and target gene expression consistency. Cell-type-specific eRegulons were ranked by descending triple_rank values to identify high-confidence, biologically relevant regulatory circuits.

### Regulation Network Analysis

For each regulatory region identified, one or more transcription factors (TFs) may bind to it, and one or more genes may be regulated by it. To quantify the similarity of eRegulons across species and brain regions, we first categorized all eRegulons by species and by anatomical brain region. For each pair of regulatory regions, we computed the Jaccard Index (JI) as a measure of similarity based on their associated transcription factors and regulated genes.

Specifically, the mathematics extension of Jaccard Index between region i and region j was defined as:

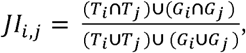

where *T_i_* and *T_j_* denote the sets of TFs associated with region i and j, respectively, and *G_i_* and *G_j_* i denote the sets of genes regulated by region i and j, respectively. This metric captures the overall regulatory similarity between two regions, incorporating both upstream regulatory inputs and downstream transcriptional outputs.

To ensure analytical robustness and reduce noise from low-confidence similarities, we applied a threshold of JI > 0.5. Pairs of regions with JI above this threshold were considered similar and connected in a resulting region similarity graph. In this graph, nodes represent individual regulatory regions, and edges represent region pairs with sufficiently high JI scores.

We defined connected components within this graph as sets of mutually similar regulatory regions—i.e., all regions within a component are connected directly or indirectly through high JI scores, and no region outside the component shares JI > 0.5 with regions within. These components represent coherent modules of regulatory regions.

For each such module, we retrieved the upstream TFs and downstream target genes for each constituent region using SCENIC+ output files. These TF–region–gene triplets were then assembled into regulatory modules, which capture putative transcriptional regulatory units conserved within or across brain regions and species.

### Measuring Difference in Homolog Cell Types

Homologous gene pairs between Homo sapiens and Mus musculus, as well as a curated list of house-keeping genes, were obtained from publicly available databases. To quantify gene expression divergence across species, we calculated a Divergence Score (DS) for each homologous gene pair within each annotated cell type (ref^33^). The DS was defined as:

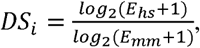

where *E_hs_* and *E_mm_* represent the normalized expression levels of the human and mouse orthologs of gene i, respectively. A pseudocount of 1 was added to avoid division by zero and to stabilize log-transformed values at low expression levels.

Genes with an absolute divergence score |DS|>2 were considered highly divergent genes, reflecting strong species-specific expression patterns. For each bio-type (Ex, Inh, Glia), we performed functional annotation of the identified divergent genes using established gene ontology (GO) enrichment tools (ref) to infer potential biological relevance and evolutionary constraints.

### Trajectory/Psudo-time Analysis

Cells annotated as Exc_L2/3IT neurons were subset from both human and mouse datasets. For each species, corresponding brain regions were merged to ensure anatomical comparability. Trajectory analysis was performed using Monocle3 (ref^48^), following the recommended pipeline. A CellDataSet (CDS) object was constructed using preprocessed expression matrices and metadata. Pseudotime values were inferred through reversed graph embedding based on the integrated UMAP coordinates.

To identify the trajectory root(s), UMAP embeddings were first visually inspected by species label to identify candidate root regions enriched for nuclei from one species. Pseudotime values were divided into 50 bins, and the relative proportion of human versus mouse nuclei within each bin was calculated after sample size normalization. This procedure enabled the assignment of biologically plausible root(s) for trajectory inference.

Pseudotime-dependent gene expression changes were identified using graph_test in Monocle3, and differentially expressed genes (DEGs) along the trajectory were clustered into co-expression modules. Module scores were calculated for each subcluster defined by MultiK (Ref), and were used as high-dimensional representations of dynamic transcriptional programs.

To quantify species-specific divergence of modules, both Euclidean distance and Manhattan distance between species-specific module centroids were computed. For each module, the centroid of expression in mouse (*G_j,mm_*) and in human (*G_j,hs_*) was calculated as:

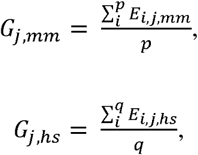

where p and q denote the number of mouse and human nuclei, respectively; *E_i,j,mm_* and *E_i,j,hs_* refer to the expression level of gene j in the i-th nucleus from mouse and human. The distances between species centroids were calculated as:

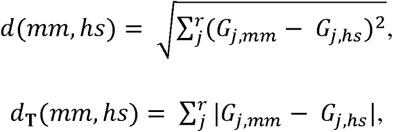

where r represents the number of homologous genes in the module. These metrics were used to assess the extent of divergence in gene expression dynamics across species along the Exc_L2/3IT trajectory.

### Integrating with PTSD GWAS data

To evaluate the contribution of regional cell types to post-traumatic stress disorder (PTSD), we utilized MAGMA (Ref) for gene set enrichment analysis. PTSD genome-wide association study (GWAS) summary statistics were obtained from Psychiatric Genomics Consortium (https://pgc.unc.edu/). SNP loci files (.bim, .bed, .fam) and the reference genome for East Asian populations were obtained from the 1000 Genomes Project (ref^54,55^) and NCBI (https://www.ncbi.nlm.nih.gov/), respectively. SNPs were annotated to genes using the MAGMA (ref^53^) annotation function with the appropriate genome build.

Gene-level association analysis was conducted using MAGMA, incorporating the PTSD GWAS summary statistics. Regional marker features were derived from both snRNA-seq (Regional Marker Genes) and snATAC-seq (Regional Marker Peaks), where marker peaks were mapped to their associated genes. These two sets of gene lists were merged per regional cell type, and duplicated gene entries within each region-specific cell type were removed to avoid redundancy. The resulting non-redundant gene sets were used as input for MAGMA’s gene set analysis.

MAGMA gene set results were visualized using lollipop plots to represent the statistical contribution of each regional cell type to PTSD genetic risk. From the .gsa.out file, we extracted key statistics, including NSNPS (number of SNPs per gene) and ZSTAT (Z-score statistics indicating association strength), for downstream annotation.

The .gsa.out file was then joined with the Regional Marker Gene list to annotate marker genes with their corresponding NSNPS and ZSTAT values, forming a GWAS-annotated gene set. Similarly, we joined this annotated table with the Regional Marker Peak list (via gene mapping) to obtain GWAS-associated statistics for the peak-derived genes, producing a GWAS-annotated peak-to-gene association table

To further assess the regulatory relevance of PTSD-associated genes, we utilized eRegulon sets from SCENIC+ analysis, including eRegulons from human and mouse. For each of these sets, target genes were used to perform an inner join with both the GWAS-annotated gene list and the GWAS-annotated peak gene list. For the peak-based analysis, consistency between eRegulon annotation (Annotation for Gene) and regional peak annotation (Annotaiton for Region) was required to ensure accurate cell-type-specific mapping.

We then investigated the distribution of NSNPS across different levels of eRegulon specificity, particularly focusing on eR_Spe_most_hs, in which both AnnoFromRegion and AnnoFromGene annotations match and are consistent with the GWAS cluster annotation. For this subset, we analyzed the composition of significantly associated genes (ZSTAT > 2), their gene ontology functions, and their roles within transcriptional regulatory networks.

### Definition and Ingestigation of Chromatin Primed Genes

To investigate the relationship between chromatin accessibility and gene expression at the regulatory level, we utilized eRegulon links derived from SCENIC+, which provide region-gene regulatory associations. Based on these associations, we evaluated the chromatin-priming status of each gene by quantifying the correspondence between chromatin accessibility and transcriptomic activity in a cell-type-specific manner.

For each region-gene pair defined in the eRegulons, we extracted chromatin accessibility values from the ATAC assay and gene expression levels from the RNA assay within the same single-cell multi-omics dataset. To systematically compare these two layers of regulation, we applied a binning strategy: both gene expression and chromatin accessibility were normalized (e.g., z-score scaled) across cells and then divided into quartiles (25%, 50%, 75%, 100%). For each cell, we counted the number of associated regions or genes falling within each quartile bin and visualized the distribution to examine coordinated regulation.

To define chromatin priming, we applied the following thresholds: 1) A region was considered “accessible” if its chromatin accessibility value exceeded a certain level across all regions in the corresponding cell type. 2) A gene was considered “lowly expressed” if its expression level fell below a certain level across all genes in the same cell type.

Based on these criteria, we constructed a binary matrix (termed the CPG matrix), where rows represent potential chromatin priming gene (CPG) pairs (i.e., region-gene pairs), and columns represent brain-region-specific major and sub-clusters. An entry of “1” indicates that the CPG is present in the corresponding sub-cluster, defined by having accessible region(s) but unexpressed gene(s), suggesting chromatin priming status.

For each cell type or sub-cluster, CPGs were further annotated with: Gene ontology and functional enrichment information, GWAS-related information, specifically NSNPS and ZSTAT values derived from previous MAGMA analyses linking genes to PTSD and PTSD human bulk-RNA-seq data in vmPFC.

## Data Availability

All data produced in the present study are available upon reasonable request to the authors

## Ackowledgement

The authors gratefully acknowledge grant support from the NSFC 82001421 (to Xiang Li), NSFC 82271556 (to Xiang Li), NSFC 82171517 (to Wei Wei). Xiang Li is surpoorted by the Major Project of Science and Technology Innovation of Hubei Province (2024BCA003). Xiang Li are supported from the medical Sci-Tech innovation platform of Zhongnan Hospital, Wuhan University. Xiang Li is supported from Climbing Project for Medical Talent of Zhongnan Hospital, Wuhan University. Wei Wei is supported by Translational Medicine and Interdisciplinary Research Joint Fund of Zhongnan Hospital of Wuhan University.

## Notes

### Competing Interest Statement

The authors have declared no competing interest.

### Author Declarations

All experiments were approved by the Institutional Review Board of Huazhong University of Science and Technology (Approval No.: 2024-S124) and performed in accordance with the principle of the Helsinki Declaration II

